# Cognitive Dysfunction in the Addictions (CDiA): A Neuron to Neighbourhood Collaborative Research Program on Executive Dysfunction and Functional Outcomes in Outpatients Seeking Treatment for Addiction

**DOI:** 10.1101/2024.08.30.24312806

**Authors:** Yuliya S. Nikolova, Anthony C. Ruocco, Daniel Felsky, Shannon Lange, Thomas D. Prevot, Erica Vieira, Daphne Voineskos, Jeffrey D. Wardell, Daniel M. Blumberger, Kevan Clifford, Ravinder Naik Dharavath, Philip Gerretsen, Ahmed N. Hassan, Sheila K. Jennings, Bernard Le Foll, Osnat Melamed, Joshua Orson, Peter Pangarov, Leanne Quigley, Cayley Russell, Kevin Shield, Matthew E. Sloan, Ashley Smoke, Victor Tang, Diana Valdes Cabrera, Wei Wang, Samantha Wells, Rajith Wickramatunga, Etienne Sibille, Lena C. Quilty, CDiA Program Study Group

**Author notes:** Lead investigators who contributed equally to the study (Yuliya S. Nikolova, Anthony C. Ruocco, Daniel Felsky, Shannon Lange, Thomas D. Prevot, Erica Vieira, Daphne Voineskos, Jeffrey D. Wardell, Etienne Sibille, Lena C. Quilty).

## Abstract

**Background:** Substance use disorders (SUDs) are pressing global public health problems. Executive functions (EFs) are prominently featured in mechanistic models of addiction. However, there remain significant gaps in our understanding of EFs in SUDs, including the dimensional relationships of EFs to underlying neural circuits, molecular biomarkers, disorder heterogeneity, and functional ability. To improve health outcomes for people with SUDs, interdisciplinary clinical, preclinical and health services research is needed to inform policies and interventions aligned with biopsychosocial models of addiction. Here, we introduce Cognitive Dysfunction in the Addictions (CDiA), an integrative team-science and translational research program, which aims to fill these knowledge gaps and facilitate research discoveries to enhance treatments for people living with SUDs.

**Methods:** The CDiA Program comprises seven complementary interdisciplinary projects that aim to progress understanding of EF in SUDs and investigate new biological treatment approaches. The projects draw on a diverse sample of adults aged 18-60 (target *N*=400) seeking treatment for addiction, who are followed prospectively over one year to identify EF domains crucial to recovery. Projects 1-3 investigate SUD symptoms, brain circuits, and blood biomarkers and their associations with both EF domains (inhibition, working memory, and set-shifting) and functional outcomes (disability, quality of life). Projects 4 and 5 evaluate interventions for addiction and their impacts on EF: a clinical trial of repetitive transcranial magnetic stimulation and a preclinical study of potential new pharmacological treatments in rodents. Project 6 links EF to healthcare utilization and is supplemented with a qualitative investigation of EF-related barriers to treatment engagement for those with substance use concerns. Project 7 uses innovative whole-person modeling to integrate the multi-modal data generated across projects, applying clustering and deep learning methods to identify patient subtypes and drive future cross-disciplinary initiatives.

**Discussion:** The CDiA program has promise to bring scientific domains together to uncover the diverse ways in which EFs are linked to SUD severity and functional recovery. These findings, supported by emerging clinical, preclinical, health service, and whole-person modeling investigations, will facilitate future discoveries about cognitive dysfunction in addiction and could enhance the future clinical care of individuals seeking treatment for SUDs.

**Scope statement:** Substance use disorders (SUDs) are a leading contributor to psychiatric morbidity, mortality, and functional impairment. Cognitive dysfunction contributes substantially to this impairment, and plays a prominent role in causal models of addiction. Executive dysfunction in particular is linked to the onset and persistence of SUDs. Large-scale, multidisciplinary efforts are necessary to elucidate the nature and course of cognitive deficits and associated functional outcomes in SUDs. The Cognitive Dysfunction in the Addictions (CDiA) integrative, translational research program undertakes to advance the study of cognitive dysfunction in SUDs, with a focus on impairments in executive function. The Program incorporates preclinical, clinical, and health systems level investigation. The proposed investigations are well positioned to determine how experimental findings translate from humans to animals (reverse translational approach), as well as how preclinical findings will translate into the development of novel treatment avenues.

## Background

Substance use disorders (SUD)^1^ affect 162 million people worldwide and are associated with substantial morbidity, mortality, and disability (1–4). The socioeconomic and health impacts associated with the use of alcohol, cannabis, and illicit drugs, substantially contribute to the global burden in years living with a disability and lost to premature death (5,6). In 2020, the World Health Organization heightened their calls for enhanced actions to curb the harmful effects of alcohol use, underscoring the global public health priority of the damaging consequences of alcohol use (7). In the US, 140,000 people die annually from excessive alcohol consumption (i.e., consuming more than 25 grams of ethanol per day) and another 70,000 from drug-related causes, costing the U.S. economy 191.6 billion dollars related to alcohol and 151.4 billion dollars due to use of other substances (8–10). Similarly, in Canada, alcohol and drug use causes almost 20,000 deaths, and costs the Canadian economy nearly 40 billion dollars annually (3). Treatments for SUDs have evidence supporting their efficacy but their availability is limited, highlighting the critical need to increase access to treatments and develop novel and improved interventions for SUD (11).

To advance the understanding of addiction, research on the cognitive and neurobiological factors involved in the development and maintenance of SUD is crucial (12). Contemporary models underscore the centrality of cognition to different phases of SUD development, with a strong emphasis on executive functions (EF) such as response inhibition and decision-making, in conjunction with motivational (e.g., incentive salience, cue reactivity/craving) and affective (e.g., negative emotionality) factors (13,14). While there is ongoing debate about the precise number of “core” processes that define EF, converging evidence suggests that EF comprises at least three factors that are related but distinct on a behavioural and neurobiological level (15–19): 1) *inhibition*, or the ability to prevent the processing of irrelevant information in working memory and/or inhibit a context-inappropriate behavioural response; 2) *working memory (updating)*, or the ability to monitor the contents of working memory for relevance to the current task and to remove from or add information to working memory; and 3) *set shifting*, reflecting the ability to switch between multiple operations or task sets. These processes are supported by common and dissociable neural substrates within a distributed network of frontoparietal brain regions (20), subserved by known biological and neurotransmitter systems (21,22), and may be partially genetically influenced (23). EF “deficits” refer to impaired functioning in one or more of these domains, whereas EF “biases” reflect the specific or selective influences of motivational or emotional contexts or materials on EF performance (24).

SUDs can be associated with both deficits and biases in EF accompanied by structural and functional alterations in underlying neural circuitry (25). Problems with EF lead to functional impairment across all stages of addiction. Interventions that bolster EF may mitigate the influence of attentional and behavioural control on drug-seeking or relapse. For example, computerized cognitive retraining may improve working memory capacity (26–28) and reduce impulsive choice and valuation of specific rewards in addiction (29). Further, non-invasive brain stimulation has exhibited promise across a range of SUDs and appears to impact both craving and decision-making processes (30–32). However, executive dysfunction in addiction has been under-investigated (33). Furthermore, the deficits and biases of EF most linked to functional outcomes remain to be identified and targeted for maximal therapeutic benefit.

Studies to date have most commonly used relatively “pure” or homogeneous patient groups (e.g., adults with a specific SUD and with no psychiatric comorbidities), with more limited attention to individual differences or within-group variability (34–36). Yet, adults seeking treatment for addiction represent a highly heterogeneous group, with complex substance use histories and a high prevalence of psychiatric comorbidities (37–39). Since “pure” group studies have limited generalizability to patient populations, there is a critical need to characterize heterogeneity of executive dysfunction in a large inclusive cohort study involving a complex patient population that is characteristic of large tertiary care facilities.

Here, we introduce the Cognitive Dysfunction in the Addictions (CDiA) Program, an integrative, team-science and translational research program focused on EF deficits and biases related to SUDs, which will aid the discovery of shared versus uniquely affected EF domains (40,41). Parallel biomarker studies in human participants and translational mechanistic studies in relevant preclinical models will link domains of EF to putative biological underpinnings, together paving the way for the rational design of targeted and individualized therapeutics for treatment and recovery from SUDs. Linkages to health care administrative databases could further mobilize knowledge to help shape public health policy and effect change at the societal level.

The CDiA Program benefits from the full participation of a Lived Expertise Research Advisory (LERA) committee, which is composed of members of the community who self-identify as having lived experience with SUD. Incorporating the perspectives of individuals with lived/living experience helps to increase the impacts of the research on the communities expected to benefit from the findings (42). In conjunction with the scientists leading the program, the LERA committee contributed to the formulation of the program and has offered ongoing insights into its public-facing materials and study procedures. The LERA committee will play a central role in interpreting the CDiA findings and sharing the results with knowledge users in the community. The CDiA Program also consults with an International Scientific Advisory Committee composed of SUD experts who provide ongoing guidance.

CDiA consists of seven interconnected projects (P1-7) with the following main objectives:

1. To identify domains of EF linked to functional outcomes in adults seeking treatment for AUD or SUD (P1).
2. To identify imaging biomarkers (MRI, fMRI) associated with domains of EF and those most predictive of functional outcomes in adult outpatients seeking treatment for AUD/SUD (P2).
3. To identify biomarkers mechanistically associated with EF and functional outcomes in adult outpatients seeking treatment for addiction (P3).
4. To assess the impact of repetitive transcranial magnetic stimulation (rTMS) on EF deficits in adult outpatients seeking treatment for comorbid alcohol use disorder (AUD) and major depressive disorder (P4).
5. A preclinical study of the impact of alcohol on EF deficits and their reversal by novel therapeutic interventions (P5).
6. To assess links between EF, treatment seeking for SUDs, and healthcare utilization and costs (P6).
7. To identify subtypes of individuals seeking addiction treatment using cross-disciplinary data types from all projects to map the biopsychosocial drivers of cognitive dysfunction in SUDs (P7).

An overview of the CDiA Program is presented in Figure 1. With the exception of the preclinical project (P5), all projects draw on a novel sample of SUD patients at the Centre for Addiction and Mental Health, allowing for the triangulation of evidence within the same pool of participants. A detailed description of each project, including the background, methods, and analytical approach, is described below.

**Figure 1.**
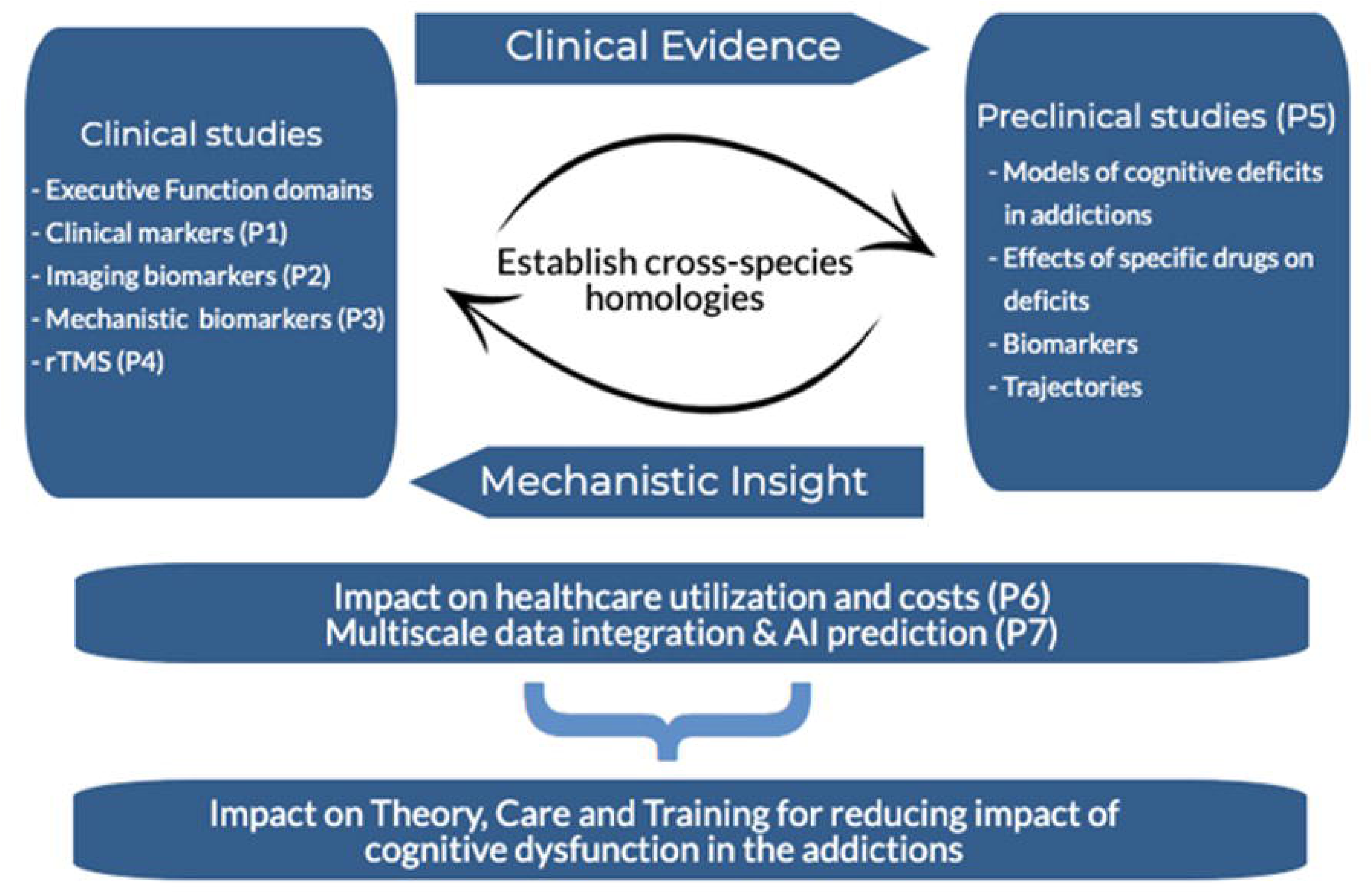

## P1: Identifying domains of EF linked to functional outcomes in adult outpatients seeking treatment for addiction

### Background

Dual Process models of addiction suggest that addiction can result from an imbalance between a controlled process that is more intentional (for which EF are central), and an automatic process that is more implicit or involuntary (for which affective states are influential) (43,44). EFs modulating either appetitive (desirable) or aversive (unpleasant) motivational states are compromised in those with SUDs ((45); also see the Addiction Neuroclinical Assessment (13,46,47)). Although many studies have investigated how deficits in EFs relate to outcomes among people with SUDs (48), the existing studies are often limited by small samples, very brief (3-6 month) follow up periods, narrow focus on a single substance in the absence of mental health comorbidities, and limited attention to functional outcomes (48). Further, most studies examine either EF deficits (i.e., general impairments in EF performance) or biases (i.e., EF impairments in the context of emotional or motivational material), without examining their relative importance for SUD outcomes. Thus, there is a need for integrative research incorporating all of these factors, and carefully evaluating links with both clinical and functional outcomes in a heterogeneous clinical sample followed prospectively over time.

### Objectives

The primary goals of P1 are to characterize EF deficits and biases and to determine which domains of EF are most predictive of functional and treatment outcomes in a complex population of adults seeking treatment for AUD/SUD in order to test the following hypotheses (H1a-e):

**<u>H1a</u>**: Deficits and biases in the three EF domains will exhibit stronger baseline associations with indicators of functioning, such as disability and quality of life, relative to other components of cognitive functioning.

**<u>H1b</u>**: EF deficits across all three domains will be most prominent in those with greater (i) severity, (ii) duration of AUD/SUD, whereas EF biases will be most prominent in those with concurrent mood disorders and elevated suicidality.

**<u>H1c</u>**: EF deficits and biases can be used to identify clusters of participants that are associated with distinct clinical features.

**<u>H1d</u>**: Relatively more severe EF deficits and biases at baseline will predict slower improvements in functional outcomes over and above substance use, treatment engagement, and healthcare utilization.

**<u>H1e</u>**: EF domains will show improvements from baseline to the one-year follow-up. Improvements in EF will be positively associated with improvements in AUD/SUD symptoms and in functional outcomes.

### Methods and Planned Analyses

#### Design and Participants

The CDiA Program aims to recruit a total sample of 400 adults seeking treatment for AUD and/or SUD to participate in this observational, longitudinal study involving clinical and cognitive assessments at baseline and over a one-year follow-up period (see Table 1 for schedule of assessments). Inclusion criteria are: (1) 18 - 60 years of age; (2) meeting diagnostic criteria for a current (past year) AUD or SUD (not counting nicotine- or caffeine-related disorders); and (3) seeking support for substance use concerns. Exclusion criteria include: (1) acute intoxication or withdrawal; (2) active psychosis; (3) acute suicidality; and (4) history of severe head injury, dementia, severe neurodevelopmental disorders, and other medical conditions or medications that could severely impair cognition.

**Table 1.** Schedule of Clinical & Cognitive Assessments.

| Procedure | Duration (min) | 0 | 2 | 4 | 6 | 8 | 10 | 12 |
| --- | --- | --- | --- | --- | --- | --- | --- | --- |
| Informed Consent | 10 | X |  |  |  |  |  |  |
| Verbal confirmation of 12 hour abstinence | 5 | X |  |  |  |  |  | X |
| Urinalysis and Breathalyzer | 5 | X |  |  |  |  |  | X |
| Interview Measures | 90 |  |  |  |  |  |  |  |
| Assessment Session Last Drug Use Verification Form |  | X |  |  |  |  |  | X |
| Timeline Follow Back (past 60 day use) |  | X | X | X | X | X | X | X |
| Contemplation ladder + Substance Use Goals |  | X | X | X | X | X | X | X |
| Diagnostic Assessment Research Tool (DART) |  | X |  |  |  |  |  | X |
| Columbia Suicide Severity Rating Scale |  | X |  |  |  |  |  | X |
| HELPS Brain Injury Screening Tool |  | X |  |  |  |  |  |  |
| Medical History |  | X |  |  |  |  |  | X |
| Psychiatric Treatment History |  | X |  |  |  |  |  | X |
| Psychological/Psychiatric Treatment |  | X |  |  |  |  |  | X |
| Self-Report Measures | 90 |  |  |  |  |  |  |  |
| Demographics |  | X |  |  |  |  |  |  |
| Bem Sex Role Inventory (Short Form) |  | X |  |  |  |  |  |  |
| Lifetime substance use history |  | X |  |  |  |  |  |  |
| Alcohol Use Disorders Identification Test |  | X |  |  |  |  |  | X |
| Drug Use Disorders Identification Test |  | X |  |  |  |  |  | X |
| Fagerström Test for Nicotine Dependence |  | X |  |  |  |  |  | X |
| Alcohol Use Awareness and Insight Scale |  | X |  |  |  |  |  | X |
| Substance Use Awareness and Insight Scale |  | X |  |  |  |  |  | X |
| Severity of Dependence Scale |  | X | X | X | X | X | X | X |
| Brief Substance Craving Scale |  | X |  |  |  |  |  | X |
| Stages of Change Readiness and Treatment Eagerness Scale |  | X |  |  |  |  |  | X |
| Substance Use Motives Measure |  | X |  |  |  |  |  | X |
| Medicinal Cannabis Use And Motives |  | X |  |  |  |  |  | X |
| WHO Disability Assessment Schedule 2.0 |  | X | X | X | X | X | X | X |
| WHO Quality of life - Brief |  | X | X | X | X | X | X | X |
| Patient Health Questionnaire |  | X |  |  |  |  |  | X |
| Generalized Anxiety Disorder |  | X |  |  |  |  |  | X |
| Childhood Trauma Questionnaire |  | X |  |  |  |  |  |  |
| List of Threatening Experiences Questionnaire |  | X |  |  |  |  |  | X |
| PTSD Checklist for DSM-5 |  | X |  |  |  |  |  | X |
| Wender Utah Rating Scale |  | X |  |  |  |  |  | X |
| Personality Inventory for DSM – Brief Form (PID-5-BF+) |  | X |  |  |  |  |  | X |
| UPPS-P Impulsivity Scales- Short Form |  | X |  |  |  |  |  | X |
| Coping Orientation to Problems Experienced Inventory, abbreviated |  | X |  |  |  |  |  | X |
| Difficulties in Emotion Regulation Scale -Short Form |  | X |  |  |  |  |  | X |
| Performance-Based Measures | 100 |  |  |  |  |  |  |  |
| CNS-Vitals |  | X |  |  |  |  |  | X |
| Executive Function Tasks: |  |  |  |  |  |  |  |  |
| Flanker Task |  | X |  |  |  |  |  | X |
| N-back Task |  | X |  |  |  |  |  | X |
| Switch Task |  | X |  |  |  |  |  | X |
| Iowa Gambling Task |  | X |  |  |  |  |  | X |
| Delay Discounting Task |  | X |  |  |  |  |  | X |
| Probability Discounting Task |  | X |  |  |  |  |  | X |

#### Measures

Interview measures include the Diagnostic Assessment Research Tool (DART; (49)) to characterize AUD/SUD and comorbid diagnoses, and the Timeline Follow-Back (TFLB; (50)) to characterize alcohol and substance use over the past 60 days. Self-report items assessing lifetime history of alcohol and substance use are also administered, along with measures of severity of alcohol- and substance-related harms. Other factors relevant to understanding substance use patterns are also assessed, including demographic factors, readiness to change, substance use motives, SUD insight and SUD treatment history (see Table 1 for a full list of measures).

A **comprehensive assessment of global cognitive functioning** is conducted with the **Central Nervous System Vital Signs** (CNS-VS; (51)) computerized assessment, which has been validated for use in addiction. CNS-VS contains 10 tests that yield 15 individual domain scores, including composite scores assessing Neurocognitive Index, Composite Memory, Psychomotor Speed, Complex Attention, and Cognitive Flexibility scores. Scores are standardized based on an age-matched normative sample. Further, tasks assessing risk taking and decision-making (**Iowa Gambling Task** (52)), and probabilistic and delay discounting (53–55) are also administered.

**EF assessment:** To achieve our goal of measuring both (a) general executive function deficits and (b) biases in these functions associated with emotional and motivational states, both neutral and emotional stimuli are used in our executive functioning tasks. The stimuli for the executive functioning tasks are a pool of happy, neutral, and sad faces from the Karolinska Emotional Directed Faces database (KEDF; (56)). Each task is administered with trial blocks with emotional stimuli (i.e., to measure executive function biases) and trial blocks with neutral stimuli (i.e., to measure executive function deficits). A **flanker task** (57–60) is used to assess **inhibition**, an **n-back task** (61,62) is used to assess working memory **updating**, and a **switch task** (63) is used to assess **set shifting**.

### Power Analysis and Data Analytic Plan

Multiple regression will be used with the clinical, functional, and demographic factors entered as simultaneous predictors of each EF domain to test hypotheses about which factors are most strongly associated with specific EF domains (H1a and H1b).

For H1c, we will conduct latent profile analyses using domains of EF as indicators of discrete latent profiles of relative deficits and biases. We will then examine the prediction of membership in these latent profiles from substance use profiles.

For H1d, latent growth curve modeling will be used to model changes in substance use frequency, AUD/SUD severity, and functional outcomes over the follow up time points. Next, rate of change will be examined as a function of baseline EF (H1a) by regressing variability in the slopes for the growth factors on baseline EF deficits and biases. These will be included as simultaneous predictors in the model to examine the specific EF domains and biases that are most predictive of change in the clinical and functional outcomes.

For H1e, we will use a generalized linear mixed effects modelling approach. EF deficits and biases will be modelled as dependent variables and time will be entered as a within-subjects factor to determine if there is significant within-person change from baseline to one year follow up. Changes over time in AUD/SUD symptoms and functional outcomes will be modelled using the same approach, with changes in EFs included in the models as a time-varying predictor of changes in these outcomes.

## P2: To identify neuroimaging biomarkers associated with domains of EF and addiction symptoms and most predictive of functional outcomes

### Background

Functional magnetic resonance imaging (fMRI) studies in healthy adults suggest that the three dimensions of EF rely on overlapping nodes within a broader frontoparietal circuit, but may also have at least partially dissociable neural substrates within and beyond this circuit (64–67). The ability to maintain a task goal stably, shared across all EF domains, relies on areas of the lateral prefrontal cortex (PFC), which extend from frontopolar regions through the mid-dorsolateral prefrontal cortex (dlPFC)(15,68,69), potentially also including the anterior cingulate cortex (ACC) and frontal operculum (70). Additional studies investigating the three EF domains side by side show overlapping activation in the inferior frontal gyrus, anterior cingulate cortex and bilateral parietal cortices (66). In contrast, behavioural inhibition is associated with more extensive recruitment of temporal regions and may engage the cortico-striatal circuit (71). Set shifting may, in turn, involve more posterior dlPFC regions (e.g., inferior frontal junction) and more extensive parietal areas (e.g., intraparietal sulcus) (72,73). Working memory updating has been additionally mapped onto fronto-striatal connections and requires input from the basal ganglia and cerebellum (74–76). Similar partially dissociable signatures of the three distinct EF dimensions have been identified in studies using resting-state (i.e., task-free) fMRI (64,65), structural MRI (77,78), and diffusion-weighted imaging (DWI)(77).

Meta-analyses of task-based fMRI across SUDs highlight the heterogeneity of paradigms used to evaluate EF, but show altered activation in fronto-parietal and striatal regions across studies (79). Importantly, however, prior studies in SUDs have almost universally focused on a single EF domain, providing relatively limited insight into the complex cognitive architecture that may underlie functional impairment in addiction (80–82). Clinically relevant insight is further limited by a predominance of cross-sectional studies, which do not investigate how neural features supporting EF may contribute to symptom change and functional recovery over time.

Characterizing executive dysfunction heterogeneities and underlying neural circuits longitudinally across large naturalistic cohorts of patients seeking treatment for SUDs is likely to provide novel insight into the pathophysiology of addiction and help identify novel targets for intervention.

### Objectives

The primary goals of CDiA Project 2 (P2) are to identify imaging biomarkers (MRI, fMRI) associated with domains of EF, their associations with addiction symptoms and day-to-day functioning, and prospective relationships with symptom changes and functional outcomes in adult outpatients seeking treatment for SUDs. These imaging biomarkers may help guide future development and individualization of biologically informed treatments for naturalistic clinical populations with SUDs.

**<u>H2a</u>**: Behavioural performance along the three EF domains will map onto partially dissociable neural substrates overlapping with frontoparietal and corticostriatal circuits. The extent of activation and pattern of brain activity associated with each of the three EFs will be associated with greater addiction severity and lower general functioning at baseline.

**<u>H2b</u>**: Greater disturbances in EF-associated neural circuits at baseline will be associated with lower improvement in addiction severity and functional outcomes on follow-up, above and beyond impairments in other neural circuits.

### Methods and Planned Analyses

#### Participants

All participants included in the total sample are eligible to participate in a 1.5-hour MRI protocol. P2-specific exclusion criteria include the presence of MRI-incompatible metal implants, history of stroke, and claustrophobia. We anticipate approximately 50% of the entire sample (n=200) will be eligible and consent to MRI. Participants are eligible for a longitudinal scan at one-year follow-up; we anticipate 25% (n=50) being available and consenting to the repeat scan to assess longitudinal stability of our main measures.

#### MRI methods

Our research imaging protocol uses state-of-the-art imaging sequences fully harmonized with MRI acquisition protocols used by large-scale population-representative studies (83) and other ongoing CAMH-based cohort studies (84,85) (see Table 2), which will allow us to leverage our data for comparison or transdiagnostic analyses. To support new research studies, a portion of the scanning time is secured for pilot sequences.

**Table 2.** MRI Sequences.

| **<u>Whole-brain MRI Sequence</u>** | **<u>Duration (minutes)</u>** | **<u>Measure</u>** | **<u>Technical Parameters</u>** |
| --- | --- | --- | --- |
| 3D T1 Magnetization Prepared RAPid Gradient Echo (MPRAGE) | 6:11 | Total, interhemispheric, and/or regional cortical thickness (mm), cortical surface area (mm<sup>2</sup>), total and regional brain volume (mm<sup>3</sup>) | Field of view (FOV) 256 x 256 mm<sup>2</sup><br>TR = 6.952 ms<br>TE = 2.920 ms<br>TI = 1060 ms<br>1 mm isotropic voxel size<br>Parallel imaging factor ARC = 2 Slices per 3D slab = 208 |
| Axial resting state functional MRI (fMRI) | 10:00 | Resting-state connectivity | FOV 216 x 216 mm<sup>2</sup><br>TR = 800 ms<br>TE = 30 ms<br>Number of slices = 60 (no gap)<br>Flip angle = 52°<br>Matrix 90 x 90<br>2.4 mm isotropic voxel size<br>Hyperband multi-slice acceleration factor = 6<br>Run time time points 750 |
| Axial N-Back fMRI | 9:40 | Executive Functioning: working memory | Same parameters as before<br>Run time 725 |
| Axial GoNoGo fMRI | 7:16 | Executive Functioning: inhibition | 545 |
| Axial Letter Judgement Task fMRI | 7:36 | Executive Functioning: set-shifting | 570 |
| Axial Diffusion MRI | 7:11 | Brain tissue microstructure | FOV 240 x 240 mm<sup>2</sup><br>TR = 4100 ms<br>TE = 81.7 ms<br>Number of slices = 81 (no gap)<br>Matrix 140 x 140<br>b-values (directions): b0 (8), b=500 s/mm<sup>2</sup> (6), b=1000 s/mm<sup>2</sup> (15), b=2000 s/mm<sup>2</sup> (15), b=3000 s/mm<sup>2</sup> (30)<br>Hyperband multi-slice acceleration factor = 3 |
| Oblique Axial Neuromelanin GRE | 8:01 | NeuroMelanin Sensitive MRI | Partial brain coverage with FOV = 165 x 220 mm<sup>2</sup><br>TR = 284 ms<br>TE = 4.1 ms<br>Number of slices = 10<br>Slice thickness = 2.5 mm<br>Slice gap = 0 mm<br>Flip angle = 50°<br>Matrix = 320 × 512 |

#### Structural measures

A standard T1-weighted structural scan is acquired to obtain detailed measures of cortical morphology (e.g., thickness, surface area, and volume). Diffusion-weighted imaging (DWI) is acquired to measure white matter tract integrity and tissue microstructure.

Neuromelanin-sensitive (NM-MRI) scans are acquired to quantify the integrity of the substantia nigra (86,87), with partial coverage of the locus coeruleus (88). NM-MRI scans were included to capture metrics related to catecholamine signaling implicated in addiction based on molecular imaging and preclinical models (89).

#### Resting-state BOLD

Participants are asked to remain awake with their eyes open for 10 minutes while lying restfully in the scanner.

#### Cognitive fMRI paradigms

We administer three EF tasks for a total duration of ∼30 minutes. The tasks assess the same EF domains as in P1 but were specifically selected to be distinct from the paradigms used in P1 to reduce practice effects, which may alter in-scanner performance and brain activation patterns (90,91). The EF cognitive paradigms were adapted from Rieck et al (66) and are presented in a pseudorandom order counterbalanced across participants. To minimize visual distraction and permit joint analysis across all EF dimensions, the tasks were designed to be visually simple and consistent across EF domains, using white or light-colored letter stimuli against a dark grey background.

#### Proposed Analysis

At baseline, task-based brain activation and brain structure will be investigated in association with SUD symptom severity and functional ability. Multivariate approaches (e.g., Partial Least Squares regression) will be used to investigate relationships between microstructure, function, and performance across the three EF tasks by identifying latent variables (LV) that capture covariance between the set of predictors (microstructure, performance) and outcome variables (functional activation). Individual brain scores reflecting the degree to which participants expressed the LV activation pattern will be correlated with addiction severity and functional ability at baseline and on follow-up, uncovering neural factors crucial for recovery. Age, sex, and primary substance of concern will be used as covariates.

Although it is challenging to provide a power estimate for studies involving complex multivariate analytic approaches in neuroimaging, our proposed sample size is consistent with recent empirically derived recommendations to enhance reproducibility in task-based fMRI research (92). These studies suggest that a sample size of n=120 is sufficient to obtain a test-retest voxel-wise correlation >0.80 across most paradigms. Thus, our proposed sample size of n=200 cross-sectionally and n=100 longitudinally, is in line with these recommendations and, to the best of our knowledge, larger than any prior fMRI study in SUDs. Further, based on standard power analysis tools, a sample size of 200 allows power of >0.80 to detect small effect sizes on the order of r=0.2 or Cohen’s d=0.2.

## P3: To identify biomarkers mechanistically associated with EF and functional outcomes in adult outpatients seeking treatment for addiction

### Background

The analysis of peripheral biomarkers has emerged as a promising approach to probe potential mechanisms or inform prognosis for SUDs (93,94). These markers are easily accessible and relatively inexpensive when compared to other methods (e.g., MRI or PET) (95,96), which highlights their potential utility in clinical settings. Establishing the relationship between peripheral biomarkers and cognitive dysfunction in SUDs can help to unravel mechanisms that contribute to SUDs, including domains of executive dysfunction, and to understand associated risk and mediating factors.

Substance use can trigger the activation of microglia and astrocytes in the brain, driving their function to a neuroinflammatory response and inducing the production of pro-inflammatory biomarkers (97,98). Several of these molecules can cross the blood-brain barrier, promoting immune activation in peripheral tissues. The persistence of this pro-inflammatory feedback loop between the brain and periphery can lead to reduced neuroplasticity, structural and functional changes in key neural circuits underlying cognitive and emotional processing, leading to executive dysfunction, physical disability, and a higher risk of SUD relapse (99).

Consistent with this, prior work supports the relationship between SUDs and peripheral markers of pro-inflammatory biological processes. For example, individuals with AUD exhibit an activation pattern characteristic of a pro-inflammatory response that persists even after alcohol withdrawal (100–102). Cocaine abstinence was also associated with higher levels of pro-inflammatory cytokines (IL-2, IL-6, and IL-17) in a sex-specific manner (103). Importantly, higher IL-6 levels have been correlated with worse EF performance (104). In individuals with opioid use disorder, higher levels of pro-inflammatory cytokines (IL-1β, IL-6) were associated with worse episodic memory performance and worse treatment response to methadone (105,106). A similar pattern of increased pro-inflammatory activation is also observed in patients with cannabis use disorder (107).

The pro-inflammatory activation observed in SUDs happens in the absence of antigens and is considered *sterile inflammation*. Sterile inflammation often occurs as a response against damaged cells and molecules. A key player of sterile inflammation is the activation of the inflammasome. The inflammasome is an intracellular multiprotein complex (e.g., the NALP3 complex) that functions as a sensor of Damage-Associated Molecule Pattern (DAMPs). It leads to the activation of proinflammatory caspases and the cleavage and release of proinflammatory cytokines, such as IL-1β and IL-18, and caspase-1.

Inflammasome activation is frequently reported in different neuropsychiatric conditions (108–110). More recently, studies have linked cytokines related to inflammasome activation, in particular IL-1β, with cognitive dysfunction in different neuropsychiatric disorders (111–117). However, there is no information about whether inflammasome activation is a potential trigger and/or contributor of cognitive impairment in general, and EFs more specifically, among adults with SUDs. The overarching goals of P3 are to investigate pro-inflammatory changes and inflammasome activation as a mechanism associated with executive dysfunction in SUDs. We will focus our analysis on peripheral biomarkers in plasma. Blood samples and extracted DNA are banked for potential future integrative analyses (see P7).

### Objectives

P3 will mainly focus on the investigation of the inflammasome in SUDs and correlate it with executive dysfunction and brain structural and functional parameters. The first objective is to investigate the impact of inflammasome activation on EF domains in SUDs. Second, we will investigate the impact of inflammasome activation on structural and functional brain parameters in SUDs (integration with P2). Results will provide insight into potentially targetable molecular mechanisms underlying EF in this naturalistic sample.

**<u>H1:</u>** Higher inflammasome activation (i.e., high expression of NALP3) will be associated with: (a) worse inhibition (b) lower working memory updating, and (c) reduced set shifting ability.

**<u>H2:</u>** Higher inflammasome activation will be associated with: (a) lower gray matter in the prefrontal cortex; (b) reduced activation of the default brain network; (c) less connectivity in neural circuits underlying EFs.

### Methods and Planned Analyses

#### Blood collection, sample preparation, and storage

P3 draws on the same participant pool recruited for P1. Following consent, blood is collected into two EDTA (8 cc) and one citrate (4.5 cc) tubes. After processing, the plasma and buffy coat aliquots are immediately stored at -80°C until analysis.

#### Laboratory analysis for plasma biomarkers

We will measure the phosphorylated and non-phosphorylated proteins isoform levels of the NALP3 complex using Western Blot. The ratio between non-phosphorylated and phosphorylated isoforms of NALP3 will be used as a proxy measure of the activation of the inflammasome pathway. Additionally, we will measure the plasma levels of the pro-inflammatory cytokines IL-1β, IL-18, and Caspase-1 using a multiplex immunoassay (bead-based multiparametric assay).

The immunoassays will be analyzed using the Luminex xMAP technology^®^ (Luminex 200). The assay has very low cross-reactivity, and this technology has a higher sensitivity to detect very low concentrations of different analytes. The intra- and inter-assay coefficient of variance for the assay is below 10% for most of the analytes measured.

#### Proposed analysis

To test the association of the biomarkers with the three EF domains and neuroimaging outcomes, we will use Pearson correlation and linear multiple regression models. The linear models can be expanded to include potential covariates, if necessary. The total sample size of 400 subjects will be sufficiently powered to detect small effect sizes of correlation coefficients of 0.11 (α=0.05, statistical power of 80%) to 0.14 (α=0.05, statistical power of 95%).

## P4: To assess the role of repetitive transcranial magnetic stimulation (rTMS) on aberrant executive function in the context of major depressive disorder in adult outpatients seeking treatment for SUD (NCT06299787)

### Background

The prefrontal cortex, although well established as an efficacious target for the treatment of major depressive disorder (MDD), has recently come into favour as a therapeutic target for AUD (118). Depressive symptoms are also highly prevalent in individuals with AUD (119–123). A number of cognitive and psychological processes stemming from the prefrontal cortex, a common treatment target for repetitive transcranial magnetic stimulation, are disrupted in both MDD and AUD. In a study by Marzuk et al (124), individuals with MDD, demonstrated poorer performance in several EF assessments. Although numerous other frontal cortical and subcortical regions are involved in MDD, the DLPFC appears to focus on executive and cognitive control of negative emotion through reappraisal and suppression strategies (125).Treatment of SUDs with repetitive transcranial magnetic stimulation (rTMS) is an extremely promising frontier (126). It has been posited that there are underlying neurobiological mechanisms shared across SUDs that involve dysfunctional cortico-striatal circuits which can be modulated with rTMS applied to the DLPFC (127). Given the role of executive dysfunction in SUD, activation of the DLPFC via rTMS may aid in modulating inhibitory behavior, salience attribution and decision-making (128). A comprehensive investigation of the domains of EF (inhibition, updating, and set shifting) associated with both MDD and AUD, has yet to be conducted and will be a major contribution of the CDiA (see P1).

Repetitive Transcranial Magnetic Stimulation (rTMS) is FDA approved for the treatment of MDD and a large body of evidence supports its safety and antidepressant efficacy (118,129,130). Theta Burst Stimulation (TBS) is an efficient form of rTMS associated with both antidepressant effects (131) and alterations in plasticity in the cortex (132). TBS involves stimulation of the cortex with triplet pulses of rTMS applied at 50 Hz (burst) and delivered every 200 msec (5Hz). TBS can be applied as continuous TBS (cTBS) as a 40 second uninterrupted burst (i.e., 600 pulses) or intermittent TBS (iTBS) as a 2 second train of bursts repeated every 8 seconds for a total of 600 pulses. Importantly, TBS takes about 1/6^th^ the time of standard rTMS, translating to a higher impact and consequently more efficient treatment at a population level.

MDD is linked to hopelessness, negative affect, and attentional biases (133,134). These features are connected with aberrant DLPFC function and are part of the negative valence domain, which includes constructs of loss, sustained threat, and frustrative non-reward (135). Rodent studies have shown that TMS-mediated increase in cortical inhibition occurs through the recruitment of dendrite-targeting GABAergic inhibitory neurons, putatively implicating Somatostatin (SST)-positive GABAergic neurons, α5-GABA_A_ and GABA_B_ receptors (to be investigated in CDiA Project 5), since these are the main inhibitory cells targeting pyramidal cell dendrites, and their function in targeted pyramidal cells is mediated by these two GABA receptor subtypes (136). The role of activating this pathway on executive function domains will be investigated in P5.

TMS combined with EEG (TMS-EEG) is a powerful method to assess GABA receptor mediated inhibition, excitability, connectivity and plasticity in non-motor cortical regions including the DLPFC. Single and paired pulse TMS-EEG studies have identified several TMS-EEG indices associated with cortical inhibitory mechanisms associated with GABA receptor mediated inhibition (137–141).

Previous studies have demonstrated that rTMS acts through mechanisms linked to GABAergic inhibitory neurotransmission in the cortex. For example, we have previously reported that rTMS increased GABA receptor mediated inhibition through rTMS and that higher frequencies (i.e., 10 and 20 Hz) produced greater inhibitory change compared to lower frequencies (i.e., 1 and 5 Hz) (142). More specifically, iTBS over the prefrontal cortex alters the N100 (143,144), which is closely associated with GABA receptor mediated inhibition (139).

We have recently demonstrated that in MDD, there are abnormalities in TMS-EEG markers of GABA_A_ and GABA_B_ receptor mediated inhibition (i.e., N45 and N100, respectively) in the DLPFC. Specifically, we reported that the N45 waveform predicted depression illness state with 80% sensitivity, 73.3% specificity, and 76.6% accuracy (area under the curve = 0.829, p < .001) (145). More recent pilot data from our group in patients with depression undergoing a 6-week rTMS course demonstrated that active rTMS changed the N45 and N100 waveforms and this change was related to HRSD-17 symptom improvement. Sham rTMS, by contrast, had no effect on either the N45 or N100 waveforms. Collectively, these results suggest that both rTMS and iTBS target neurophysiological markers of GABA receptor mediated inhibition and this mechanism may be related to MDD treatment response. However, there have not been specific neurophysiological investigations into executive functions and depressive symptomatology in the context of MDD comorbid with AUD. This proposal serves as a unique window into the neurophysiology of this at-risk population, and will further investigate the correlation among rTMS treatments, indices of GABAergic inhibition and mode defined domains of executive function (inhibition, updating, and set shifting).

An emerging neurophysiological index of prefrontal cortical activity underlying executive functioning is theta-gamma coupling (TGC), which measures the degree to which high-frequency gamma oscillations are modulated by low-frequency theta oscillations (146) TGC is thought to underlie working memory, the brain’s ability to select, maintain, and manipulate information in the short-term. In a group of patients with MDD, Noda et al. (147) showed that resting state TGC levels correlated with executive function and was enhanced after 10 sessions of DLPFC rTMS, suggesting that it may be a biomarker for intervention in treatments that target brain regions underlying executive dysfunction. To further investigate TGC as a biological mechanism in MDD, the present project adds an investigation of TGC coupled with the N-back working memory task before and after rTMS treatment. The project bridges gaps in previous studies (147) by including a sham control group, measurement of TGC that is coupled with a working memory task, investigating effects in comorbid AUD, and delivering a full 20 session course of rTMS.

### Objectives

P4 will enhance the development of TBS as a new intervention for AUD in the context of depressive symptoms and uses integrated TMS-EEG to identify neurophysiological targets of executive dysfunction in this disorder. The study aims to conduct a double-blind randomized pilot study, consisting of two arms (bilateral TBS and sham) to investigate the effect of bilateral TBS on deficits in executive function in the context of comorbid AUD and MDD. We will leverage our expertise in neurophysiological investigations of the cortex through TMS-EEG to index GABA inhibition in the cortex of individuals with a diagnosis of AUD, who have not used substances within the past month (ie. meeting criteria for early remission), in the context of a current depressive episode and MDD, and index alterations in TMS-EEG indexed GABA receptor mediated inhibition induced by bilateral TBS or sham applied to the DLPFC. We will also determine whether bilateral rTMS to the DLPFC changes TGC and is associated with changes in executive dysfunction in patients with MDD and AUD. We will also assess the direct associations between improvement in executive function domains after bilateral TBS, with improvement in MDD symptoms and conduct exploratory subanalyses by biological sex and age categories. Finally, we will aggregate data from other CDiA projects (including imaging, genetics, inflammatory models, preclinical biomarkers and more) to further examine the role of GABA receptor mediated inhibition in the cognitive dysfunction found in SUD. Specific baseline cross-analyses will be performed between executive function tasks, imaging variables, inflammatory markers and TMS-EEG indices of GABA receptor mediated inhibition.

**<u>H1:</u>** 4 weeks of daily bilateral DLPFC TBS will lead to greater improvement in executive function tasks, compared to sham TBS.

**<u>H2:</u>** 4 weeks of daily active bilateral DLPFC TBS will lead to a greater proportion of individuals remaining abstinent from substance use compared to sham rTMS.

**<u>H3:</u>** 4 weeks of daily active bilateral DLPFC TBS will lead to greater amelioration of depressive symptoms (as indexed by decrease in HRSD-17 score) and suicidal ideation (as indexed by a decrease in Columbia-Suicide Severity Rating Scale score), compared to sham rTMS.

**<u>H4:</u>** Active bilateral TBS DLPFC stimulation (compared to sham) will lead to an increase in GABA receptor mediated inhibition indices after the course of TBS.

**<u>H5:</u>** Active bilateral DLPFC rTMS (compared to sham) will lead to a significantly greater increase in TGC and correlate with improvements in executive dysfunction.

**<u>H6 (exploratory)</u>**: 4 weeks of daily active bilateral DLPFC TBS (compared to sham) will lead to improvement in all executive function domains (inhibition, updating, and set shifting) in conjunction with improvement in MDD symptoms (ie. more amelioration of MDD symptoms will be associated with improved executive function).

### Methods and Planned Analyses

A total of 40 participants will be recruited from the CDiA pooled sample within 6 to 12 weeks of the baseline assessment for P1. Participants will be randomized in a 1:1 ratio to one of two different rTMS arms. The first arm will include bilateral TBS, applied as cTBS over the right DLPFC followed by iTBS applied over the left DLPFC. The second arm will be sham TBS. All participants will be asked to participate in 4 weeks of treatment, 5 days/week (i.e. weekdays).

TMS-EEG will occur at treatment initiation (week 0) and treatment end (week 4). Baseline and post-treatment clinical measures will be administered to characterize each AUD patient. TBS will be administered using the MagPro X100 stimulator equipped with a Cool-B70 coil and Qooler fluid-cooling device (MagVenture, Farum, Denmark) positioned under MRI guidance using the Brainsight neuronavigation system. With a sample size of 20 participants per group and assumed large effect sizes, we will have 80% of statistical power to detect a significant difference between the active treatment arm and sham at a significance level of 0.05.

## Randomization and Blinding

We will randomize participants based on a stratified randomization scheme using a permuted block method with a random number generator, in fixed random sizes.

## Active/Sham rTMS Apparatus/Blinding Procedure

We will use an R30 or X100 with a cool A/P B70-type coil (Magventure Inc.) to ensure blinding of both patient and technician. As such, the clinician, researcher, patient and technician will all be blinded. For both active and sham stimulation, the coil is positioned under MRI guidance using real-time neuronavigation, thus providing technician and participant with visual feedback throughout stimulation.

## TBS Treatment Procedure

TBS will be delivered to the DLPFC bilaterally, with neuronavigation determined through structural MRI. The structural MRI utilized to derive the brain target will have been completed as part of P2. Should participants not have had imaging performed as yet when they are screened for this project, they will be redirected through Project 3 after baseline assessments of this project prior to the start of rTMS treatment. The coil position selection will start by identifying MNI152 stereotaxic coordinates (x, y, z) of (–38, +44, +26) and (38, 44, 26) in the left and right DLPFC, respectively, which we have shown to be effective targets for rTMS in alleviating symptoms of MDD (148). Bilateral TBS will be delivered as follows: R-DLPFC cTBS: 40s uninterrupted bursts (triplet 50 Hz bursts, repeated at 5 Hz, 40s on, 600 pulses total) followed immediately by L-DLPFC iTBS: (triplet 50 Hz bursts, repeated at 5 Hz, 2s on and 8s off, 600 pulses total). For intensity of stimulation we will use 120% RMT. Our work in over 200 people stimulating PFC suggests that 120% RMT is the maximum that should be used for negligible risk of seizure and other serious adverse effects beyond the expected scalp pain and headache during early treatment sessions (149,150).

## Participants

Patients will be included if they meet general inclusion criteria for CDiA and in addition, if they: (1) have a Diagnostic and Statistical Manual for Mental Disorders, 5^th^ edition (DSM–5) diagnosis of AUD based on the MINI; (2) do not exhibit problematic use of any substances (excluding nicotine and caffeine), including alcohol, for ≥1 month; (3) screened positive for an MDE based on the MINI without psychotic symptoms (4) are agreeable to keeping their current antidepressant medications and medications for AUD constant during the study; (5) are reliably taking SUD agonist therapies if appropriate and managed by their clinical team; (6) are able to adhere to the study schedule (7) meet the TMS safety criteria (151).

Participants are excluded if they meet general exclusion criteria for CDiA, and in addition, if they: (1) have a concomitant major unstable medical illness or any significant neurological disorder; (2) are pregnant or intend to get pregnant during the study; (3) have failed a course of ECT, due to the lower likelihood of response to rTMS; (4) have an intracranial implant (e.g., aneurysm clips, shunts, cochlear implants) or any other metal object within or near the head, excluding the mouth, that cannot be safely removed; (5) require a benzodiazepine with a dose equivalent to lorazepam 2 mg/day or higher (152).

Participants will be discontinued from the study if they cannot safely continue the study based on any of the following criteria: (1) experience clinically significant worsening of suicidality that requires an involuntary inpatient hospitalization; (2) develop clinically significant hypomanic or manic symptoms; (3) relapse into substance use in the month prior to rTMS treatment or during the rTMS treatment (4) miss four rTMS treatments (i.e., 20%); or (5) withdraw consent.

If relapse into substance use is suspected during the course of rTMS treatment, for safety purposes, a physician will be called to assess the participant prior to the initiation of rTMS treatment for the day or as soon as substance use is suspected. Investigations (e.g., urine drug screen or blood draw) will be ordered at the physician’s discretion, with the consent of the participant. The decision to continue with or discontinue rTMS treatment for that day or going forward will be at the discretion of the assessing physician and the participant’s SUD clinical team.

## Clinical Assessments

Participants will be initially screened with the MINI and C-SSRS as part of P1. Interested and potentially eligible participants will complete a subsequent interview to address all criteria outlined above to confirm eligibility..

The **17-item Hamilton Rating Scale for Depression (HRSD-17)** (153) will be our tertiary clinical outcome measure. The tertiary clinical outcome criteria will be a decrease in HRSD-17 score at treatment end compared to baseline (129,148). The HRSD-17 will be performed at baseline, weekly and at the end of the rTMS treatment course.

The **Columbia-Suicide Severity Rating Scale (C-SSRS)** will be used to evaluate suicidality. This reliable and valid scale has been used in randomized clinical trials and is able to predict completed suicide (154). The C-SSRS has been reported as an effective measure for diagnosis and treatment across several diagnoses (155–157) with high internal consistency, and interrater reliability (157–161). The C-SSRS will be performed at baseline, weekly and at the end of the rTMS treatment course.

**TMS Adult Safety Screen (TASS)** (151) will be used at recruitment and baseline to ensure that participants are safe to participate in rTMS treatment trials.

**Executive Function Battery** To enhance integration of each project, we will re-administer the EF cognitive assessments within one week prior to the start of rTMS and at the end of the 4 week course of rTMS.

## Neurophysiological Indices (TMS-EEG) of GABA Receptor Mediated Inhibition in the DLPFC

To evaluate GABA receptor mediated inhibition indices in the DLPFC, TMS will be administered to the left DLPFC using two Magstim-200 stimulators (Magstim Company Ltd., UK) connected via a Bistim module and electrophysiological data will be collected using dedicated hardware and software (Neuroscan, Compumedics, USA). Each TMS session will include the establishment of the individual threshold for stimulation, followed by GABA receptor mediated inhibition paradigms in the DLPFC according to our published methods (139). Recordings will be acquired through a 64-channel EEG. GABA receptor mediated inhibition indices from the DLPFC will be the dependent variables of interest and derived according to published recommendations (139,162). Our neurophysiological measures have been established in several of our previous reports and have a high test-retest reliability (i.e., ICC > than 0.9) (163,164). Data analysis will take place using semi-automated methods developed and validated by our group (139,163). TMS-EEG will be conducted prior to the start of the course of rTMS and within 48 hours after the last rTMS treatment.

To evaluate TGC, participants will undergo the N-back task during a 10 minute EEG recording at baseline and after the treatment course as part of the TMS-EEG sessions. The N-back task assesses working memory updating by asking participants to determine whether the stimulus (a letter) presented to them is the same as, or different than, the stimulus that was presented to them N trials previously. Participants indicate whether the stimulus presented on the monitor is a similar match, or different, from the stimulus presented 2 trials back. EEG signals of TGC are recorded using DC and a low pass filter of 100 Hz at 20-kHz sampling rate, and data processed offline using MATLAB and EEGLAB toolbox, following previously published protocols (146,165). *<u>Theta-Gamma Coupling</u>:* The measure of TGC is indexed by the modulation index (MI), calculated for each electrode, followed by an average across the right and left frontal electrodes. The MI for all target trials is analyzed as a weighted average based on the number of correct and incorrect responses, as previously described (146,165).

## P5: A preclinical study of the impact of alcohol on executive dysfunctions (ED) and their reversal by novel therapeutic interventions

### Background

Preclinical research on substance administration played a critical role in understanding the nature of substance-related cognitive dysfunction. However, studies focused on single cognitive domains, and relatively little is known about the dynamic trajectories of executive dysfunction (i.e. emergence, dependence and potential reversal (166,167)). This study employs a reverse-translational approach, using homologous behavioral tests applied to human participants, but here in rats, to measure executive dysfunction in three domains affected by chronic alcohol consumption (Objective 1), potential reversal via pharmacological intervention (Objective 2), and identification of peripheral and central biomarkers for translation to humans (Objective 3).

The investigation targets specific biological pathways implicated in executive dysfunction in AUD, focusing on GABAergic neurotransmission, the inflammasome system, the noradrenergic system, and the opioid system. The GABAergic system plays a critical role in cognitive functions (168), and is being investigated in P4. Here, we will enhance its activity pharmacologically in rats to investigate its impact on executive functions. The activity of the inflammasome, a multiprotein intracellular complex that detects internal biological stressors and activates pro-inflammatory cytokines, is implicated in brain disorders (169) and alcohol consumption (170), and will be measured in P3 for its translational biomarker value. Here, we will block its activity pharmacologically to assess the impact on executive functions. The integrity of the noradrenergic system, to be measured in P2, plays a critical modulatory role in AUDs (171), with complex effects depending on activity at various receptor subtypes (α1 versus α2). Here we propose to target the α2-receptor, using guanfacine, an α2-receptor agonist that demonstrated efficacy at reducing alcohol consumption in rats (172) and is used for the treatment of attention-deficit/hyperactivity disorder (173). Finally, naltrexone is a prescribed medication used to manage alcohol dependence, especially consumption, although it appears to have reduced efficacy in certain patients, depending on gender and genetic differences (174). Naltrexone is predicted to be prescribed in a significant proportion of P1 participants. Its effects on executive functions are not fully characterized and tend to vary from study to study (175,176). Here, we propose to systematically investigate the relationship between its known clinical efficacy on consumption and potential improvement of executive functions in rats, providing a clinically-relevant comparison to the other interventions.

### Objectives

The primary objective of P5 will be to characterize the impact of alcohol on three cognitive domains in rats, identify the impact of novel therapeutic approaches, and delineate novel biomarkers predictive of cognitive deficit emergence or treatment efficacy. We will test the following hypotheses:

**<u>H1</u>**: Alcohol withdrawal increases working memory deficits, impulsivity and cognitive flexibility deficits in adult rats.

**<u>H2:</u>** Cognitive dysfunctions observed are linked to changes in central biomarker of synaptic functions, changes in various neurotransmitter related activity (glutamate, GABA, dopamine) and peripheral signs of neuroinflammation.

**<u>H3a</u>**: Pharmacological interventions targeting the GABAergic system will contribute to better working memory function, cognitive flexibility and reduced impulsivity during withdrawal.

**<u>H3b</u>**: Pharmacological interventions targeting the dopaminergic system will contribute to better cognitive functions and reduced alcohol seeking behaviors.

**<u>H3c:</u>** Pharmacological interventions targeting the inflammasome will reduce signs of neuroinflammation, indirectly contributing to better cognitive health.

This project will also generate a sample repository (brain, blood, plasma) for investigation and discovery of peripheral and central biomarkers indexing putative mechanisms involved in AUD and executive dysfunction, that could be used for follow-up biomarker analyses.

### Methods and Planned Analyses

Male and female Long Evans rats, 3 months old at study initiation, will be utilized in this study. Rats will be habituated to consume a liquid diet (Nestlé® Chocolate Protein Boost) in which ethanol can be diluted. After baseline assessment of executive functions, half of each cohort will be exposed to a diet supplemented with 10% w/v alcohol 5 days per week. Removal on 2 consecutive days per week will induce a withdrawal period. Such cycle will be repeated 4 times, with assessment of cognitive functions during withdrawal after 4 cycles (Figure 4; top panel, orange box)

**Figure 2.**
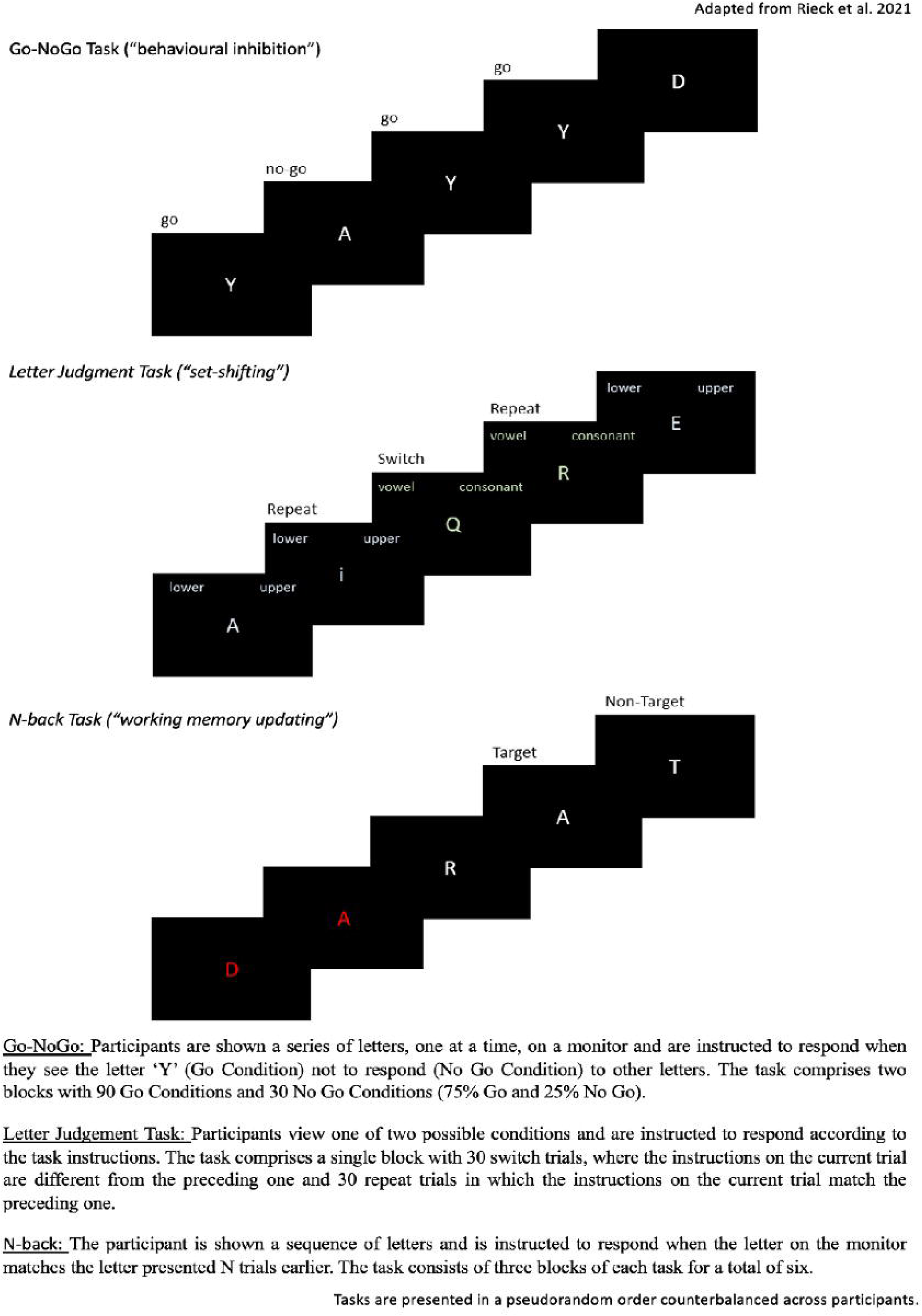

**Figure 3.**
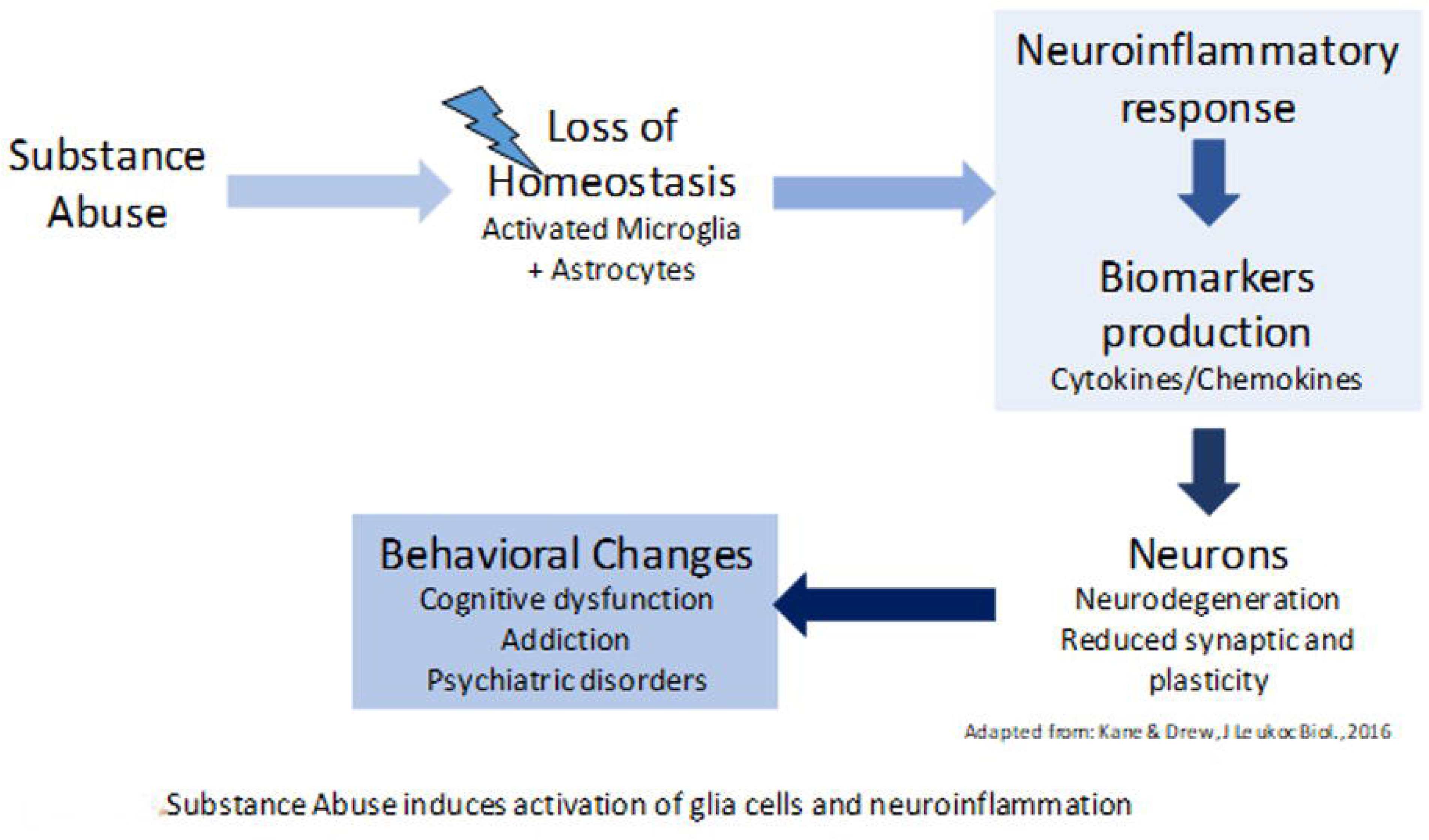

**Figure 4.**
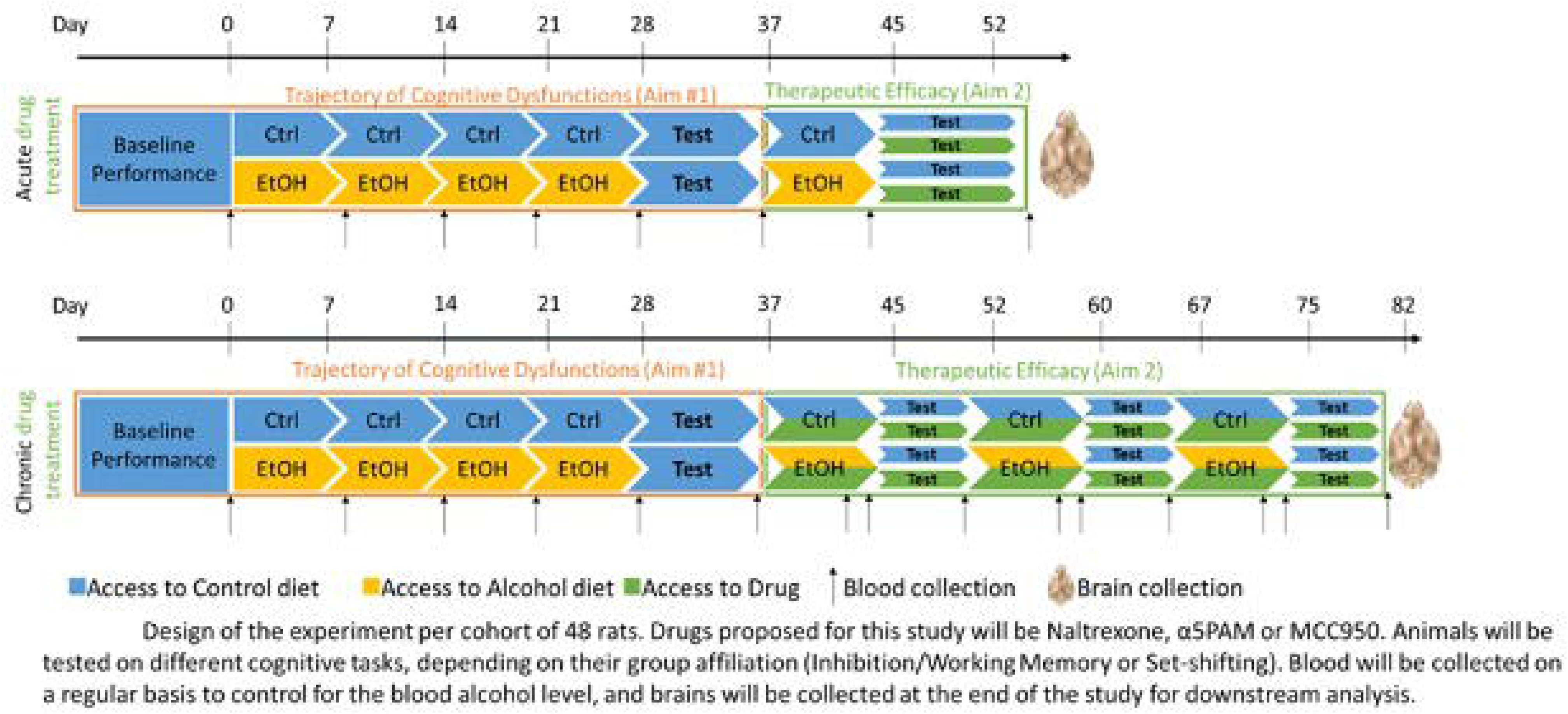

#### Measures

Three domains of EF (inhibition, working memory, set-shifting) will be assessed to match assessment in P1. **Inhibition** will be measured using the 5-Choice Serial Reaction Time Task (5-CSRTT). This test is conducted in operant conditioning boxes with 5 apertures featuring integrated light stimuli (177), and a reward dispenser on the opposite wall. Rats are trained to respond to a stimulus among the 5 apertures, earning a reward on a fixed intertrial interval.

Lengthening this interval challenges rats to wait longer before responding, with premature responses during the intertrial interval serving as a reliable measure of response inhibition (or impulsivity). In alcohol-exposed rats, premature responses are expected to increase, especially when the intertrial interval is extended. **Working memory** will be assessed using a T-maze spontaneous alternation task (178). After 2 days of habituation, rats explore the maze starting in the maze’s initial arm and then choosing between the two goal arms. The chosen arm is closed off, and the rat returns to the start arm for the next trial. This process is repeated for seven trials. Rats naturally alternate between arms, relying on working memory for novel environment exploration. Alcohol-exposed rats tested during withdrawal are expected to show diminished working memory functions **Set-shifting** will be evaluated following Floresco et al.’s protocol (179), in which rats undergo initial training in standard operant conditioning chambers with a house light, two retractable levers, and stimulus lights. Training involves a fixed ratio 1 schedule on each lever, reaching a criterion of 50 rewards or 30 minutes per lever. The second phase introduces a 90-trial session where rats must press a lever within 10 seconds to receive a reward. Visual cue discrimination training follows, with stimulus lights above the levers. Rats respond within 10 seconds to the lit lever for a sucrose reward, while responses on the non-lit lever are punished. Criterion: 10 consecutive correct responses for 30 trials over 4 days. After visual discrimination is learned, rats are tested in a “shift to response discrimination” task, where the rewarded lever is consistent, irrespective of the light stimulus. The number of trials needed to learn this new set serves as a proxy for set-shifting. Rules are modified after each testing session to ensure continual adaptation.

Due to the complexity of the inhibition and set-shifting task, these cannot be run in the same animals. Therefore, different animals will be used to run inhibition+working memory, and set-shifting.

Following the assessment of alcohol-induced cognitive function changes, interventions will commence (Objective #2). Treatments involve intraperitoneal administration of drugs targeting α5-GABAA receptors (GL-II-73), the inflammasome (MCC950), the α2-noradrenergic receptor (guanfacine), and the opioid system (naltrexone).

Initially, acute dosing will be explored using a Latin square design to test multiple doses (Table 3). Conducting multiple doses in the same animal necessitates several testing sessions, and all employed tests, such as the Y maze and 5-CSRTT, are stable and unaffected by practice and repetition (177,180–182). The set-shifting task (179) can be adapted to establish new rules at each testing session, enabling regular assessment of set-shifting abilities in the same animals while mitigating the impact of practice on repeated testing.

**Table 3.**

| **Mg/kg** | **Low dose** | **Medium dose** | **High dose** |
| --- | --- | --- | --- |
| Naltrexone | 0.1 | 0.3 | 1 |
| α5PAM | 3 | 10 | 30 |
| MCC950 | 1 | 3 | 10 |

Blood samples will be drawn weekly and 24 hours after discontinuation of the liquid diet (Figure 4) to verify 1) blood alcohol levels (BALs) exceeding 100mg% during exposure and 2) the absence of alcohol 24 hours after cessation when behavioral testing begins. BALs will be assessed using an ANALOX machine. Serum samples will also be utilized for measuring peripheral inflammasome biomarkers. Inflammasome activity will be determined by the phosphorylated/non-phosphorylated NALP3 ratio in peripheral blood mononuclear cells, along with investigating inflammatory markers (NF-ΚB, ERK, p38MAPK) and cytokines (IL-1β, IL-18, Caspase-1). Luminex xMAP technology® will be employed for biomarker analyses.

After behavioral assessments, rats will be euthanized, and brains dissected for immunofluorescence staining and molecular analyses on regions of interest (prefrontal cortex, VTA, nucleus accumbens and hippocampus). Rostro-caudal coronal sections will be stored at - 20°C for neuroinflammation, noradrenergic, and GABAergic marker analysis. RNA and proteins will be extracted from homogenized tissue, allowing for cDNA conversion, quantitative PCR, and Western Blot analyses. The markers examined include, but are not limited to, SST, PV, α5-GABRA, α1 GABRA, BDNF, and other GABAergic markers. Similarly, qPCR, immunostaining, and Western Blot analyses against sterile inflammation biomarkers (NALP3, NF-ΚB, ERK, p38MAPK, IL-1β, IL-18, Caspase-1) will validate the correlation of peripheral and central biomarkers in a region-specific manner. Investigation of changes in noradrenergic system markers (NET, α1 receptor, α2 receptor) will complement NM-MRI work outlined in P2.

The blood and brain samples collected will form a repository for subsequent biomarker analyses related to executive dysfunction in AUD. As Objective #3 establishes this repository, the analysis of these samples (peripheral or central) will be influenced by findings from other projects. Consequently, additional exploratory investigations of biomarkers will be conducted in these samples when other projects identify relevant pathways or markers in the clinical population.

#### Proposed Analysis

Behavioral data will be examined to assess the impact of alcohol exposure on executive functions, utilizing ANOVA (or relevant non-parametric alternatives) followed by Tukey’s post hoc comparisons for statistical significance. In the alcohol-exposed groups, it is possible to observe subgroups of animals with higher susceptibility to alcohol than the others, suggesting existence of high-responders and low-responders. For data collected weekly, data will be analyzed with repeated measures to identify animals that are more than 50% of the time over the mean, and animals that are more than 50% of the time under the mean to segregate high-responders and low-responders respectively. If such separation does not exist consistently, then analyses will focus on overall alcohol effect. For the second objective involving drug interactions, separate 2-way ANOVA with diet and treatment as factors will be applied, followed by Dunnett’s post-hoc comparisons (182). In Objective #3, blood sample data will be analyzed using Student’s t-test (Objective #1) or 2-way ANOVA (Objective #2) with diet and treatment as factors, followed by Dunnett’s post-hoc test. Brain analyses with qPCR and Western Blot will use 2-way ANOVA with diet and treatment as factors, followed by Dunnett’s post-hoc comparisons. Pearson or Spearman analyses will evaluate correlations between executive dysfunction and peripheral/central biomarker levels, with p-values adjusted for multiple comparisons using the Bonferroni method due to the potential high number of pairwise analyses.

## P6: To assess links between EF and treatment seeking for addiction and healthcare utilization and costs

### Background

Among individuals with a SUD, EF can affect retention (183–185) and engagement in treatment as well as treatment outcomes (37,186), including the ability to achieve and maintain abstinence (183–185,187). Given that SUDs are strongly associated with health care utilization (39), it is also likely that EF, which impacts the success of SUD treatment, is a key factor affecting the extent of health care utilization. In particular, among those with EF deficits, a diminished ability to achieve abstinence or low-risk alcohol use may lead to a higher use of emergency services for the acute consequences associated with SUDs (188). This could result in increased health care costs (189), which is problematic when existing resources dedicated to emergency services are limited (189,190). Furthermore, a diminished ability to achieve abstinence or low-risk substance use may also lead to a higher use of non-emergency health care services, including primary care visits and hospitalizations, due to acute and chronic health effects associated with substance use (191,192).

Furthermore, previous research has shown that EF is a factor in engagement in SUD treatment, including more treatment episodes and longer durations of abstinence among those without executive function deficits (187). EF deficits lead to worse recognition of problem use (193–195). Therefore, people with executive function deficits are more likely to view their symptoms as temporary or not serious, and consequently do not seek treatment (38). These factors may also affect whether patients stay in treatment or remain abstinent after treatment. Accordingly, it is likely that executive function deficits affect treatment seeking behaviours, experiences, and barriers among people with SUDs.

### Objectives

P6 has two objectives. The first is to qualitatively assess the potential impact of EF on treatment-seeking behaviors, experiences, and barriers, specifically among individuals with AUD. The second objective is to quantitatively assess the association of executive function with health care utilization among individuals with SUD, and to estimate the costs of this utilization.

**<u>H1:</u>** EF deficits will negatively impact treatment seeking and the treatment process as expressed by participants in treatment for AUD.

**<u>H2:</u>** Emergency and non-emergency health care utilization and related costs will be higher among participants in treatment for SUD with more severe EF deficits than among those with less severe EF deficits.

### Methods and Planned Analyses

#### Objective 1 (Qualitative component)

A semi-structured, one-on-one 90-minute interview with a subset of participants recruited from the pooled CDiA sample will be conducted by a research coordinator trained in qualitative methods including interviewing and analysis, shortly after their baseline data are collected. Given differences in treatment pathways and trajectories and their potential impacts on treatment experiences, this analysis will be limited to participants with AUD. Interview questions will be related to treatment-seeking behaviors and executive functioning. Participants will be asked about their experiences seeking help for substance use, perceived facilitators and barriers to treatment, perceptions and expectations regarding treatment, past health care utilization, and questions related to how executive function (e.g., memory, attention, impulsivity, etc.) may have played a potential role in their treatment-seeking behavior and experiences.

This study will undertake a thematic analysis of the data obtained, examining and identifying common themes that emerge related to the perceived impact of executive function on treatment-seeking behaviors, experiences, and barriers across the sample. We expect that thematic data saturation will be met with a sample size of approximately 30 participants.

All interview transcripts will be manually reviewed by a member of the research team, and an initial codebook will be prepared based on preliminary analyses. All transcripts will be coded for the initial set of codebook themes using NVivo software. As we anticipate both inductive and deductive themes to emerge from the data, the codebook will be subject to further development and revision based on ongoing analyses and discussion of emergent themes among members of the research team. Data analysis will be finalized once we have met data saturation, and no new themes emerge. Final results will be informed by common themes that emerged and were expressed by multiple participants. To increase data analysis credibility, a secondary independent coder will code a sub-sample of transcripts using the coding framework to assess inter-coder reliability, and any discrepancies between codes or themes will be discussed.

Subsequent to the interview, participants will be asked to complete the Behavior Rating Inventory for Executive Functioning in Adults (BRIEF-A), a standardized validated assessment that captures self-reported perceptions of an individual’s executive functions and self-regulation (196). The results of the BRIEF-A assessment tool will be used to better understand the participants’ executive functioning from their perspective, which will further inform our qualitative analysis.

#### Objective 2 (Quantitative component)

A health systems evaluation will be conducted to assess the association between EF (as measured in P1) and health care utilization, as well as the costs of this utilization. For this analysis, the database constructed in P1 will be linked with data on health care utilization and the associated costs obtained from the Institute for Clinical Evaluative Sciences (ICES). ICES will provide data on the use of publicly-funded health services in Ontario. Participant data will be linked through ICES to the Hospital Discharge Abstract Database (DAD), National Ambulatory Care Reporting System (NACRS), Continuing Care Reporting System (CCRS), Ontario Health Insurance Plan Claims Database (OHIP), and Ontario Drug Benefit Claims (ODB) database. Records-level ICES data linkages will be performed using each participant’s unique Ontario Health Insurance Program health card number; the resulting database will be stripped of all direct personal identifiers and each entry will be assigned a confidential code number. All data linkages will be performed by an ICES data analyst. A subanalysis will be performed stratifying by conditions fully attributable and those not fully attributable to SUD.

Analyses will focus on the occurrence of service utilization, stratified by the type of service (i.e., non-emergency services and emergency services). The occurrence of health care service use among the cohort (recurrent time-to-event data) will be analyzed using multiple models, including covariates across multi-disciplinary domains. These models will include the Andersen and Gill model (i.e., generalizes the Cox model), Prentice, Williams and Peterson (PWP) model, marginal means/rates model, and frailty model (i.e., random effects model) (197–200). The final model to be used will be chosen based upon which of the previously used models best describes the data and which model assumptions are upheld. Model assumptions include the occurrence of an event being dependent upon the prior number of events during the follow-up period (PWP), there being no time-dependent covariates (marginal means/rates model), and the occurrence of an event cannot be explained by observed covariates alone (frailty model). Each statistical model will include covariates for the domains of EF, demographics, substance use, and health service utilization prior to study participation.

## P7: To identify subtypes of individuals seeking addiction treatment using cross-disciplinary data types from all projects to and map the biopsychosocial all-cause drivers of cognitive dysfunction in adult outpatients seeking treatment for AUD/SUD

### Background

In SUDs, heterogeneity is the rule rather than the exception (201). This heterogeneity is largely driven by extremely high rates of comorbid psychiatric conditions (202–204) and compounded challenges in the diagnostic process, as emergent cognitive syndromes may be temporary, drug induced, or chronic (205,206). Even when considering the same SUD, symptoms and prognoses vary wildly between patients due to complex combinations of life experience (207), biology (208), and socio-demographic factors (209,210). Given this heterogeneity, it is likely that there exist subpopulations, or subtypes (211–213), of individuals with similar diagnostic classifications but different underlying disease mechanisms and degrees of cognitive impairment. Due to these mechanistic differences, interventions aimed at improving deficits in executive function (214) would not be expected to work equally well in different subtypes, meaning their identification is a priority for future applications of precision medicine in SUDs.

Attempts to define subgroups within and across diagnoses with relatively homogeneous symptom profiles and outcomes have met with limited success; for example, a five-biotype model of alcohol dependence has been proposed using latent class modeling of family history, age at onset, DSM-IV criteria, and data on comorbid illnesses (215). Others have proposed four-and two-group subtyping schemes based on psychiatric co-morbidity, family history, psychopathy, temperament, and other clinical health measures (216). Despite the strong links between symptom dimensions and executive dysfunction (see P1), subtypes have not been formally evaluated for differences in domains of inhibition, set shifting, or working memory updating, leaving a major gap in the field. To their detriment, most efforts have focused on relatively few domains of interest, not examined changes in cognitive performance (specifically executive function) over time, and yielded varying, unstable results. Therefore, it is important to approach the problem from a data-inclusive perspective (217). Using insights from unimodal investigations, such as those proposed in the preceding CDiA projects, it is also possible to assemble optimal parsimonious predictive models at the individual level which could aid in screening efforts and clinical decision making.

### Objectives

In this project, we propose to integrate data collected from each of the proceeding projects – including use of insights from animal experiments in P5 – to (1) perform multi-modal subtyping on SUD individuals, (2) evaluate subtypes against known biological risk profiles for cognitive dysfunction and psychiatric illness (such as exploring polygenic risk scores) and other longitudinal outcomes (e.g. TMS treatment outcome, specified in P4), and (3) use machine learning approaches to optimize predictors of cognitive dysfunction over time and to understand the structure of modifiable risk and resilience factors that may present opportunities for intervention. No additional data will be collected for this project; it will be computational in nature.

**<u>H1:</u>** Multi-modal unsupervised and semi-supervised clustering of SUD patients at baseline will reveal transdiagnostic subtypes of SUD with unique risk factor profiles and levels of executive functioning.

**<u>H2:</u>** Subtypes from Aim 1 will be predictive of changes in executive function over one year of follow-up and individuals with evidence for subgroup transition will experience different treatment response rates.

**<u>H3:</u>** Multivariate predictive models of changes in executive function over time, incorporating the available multi-modal feature space of CDiA, will offer better performance than models based on individual modalities.

### Methods and Planned Analyses

#### Cross-sectional data-driven subtyping

This will involve using subject clustering algorithms to identify relatively homogeneous subgroups of patients. Primary measurements from each of P1-6 will be used as input for clustering (see **Table 4** for domain definitions). The similarity network fusion (SNF) method will then be used to combine more fine-detailed, high-dimensional input variables across multiple modalities. To address the key overarching theme of this proposal, subgroups based on executive function domains (including and excluding other measures) will be of particular interest, and will be tested for distributional differences in primary prognostic outcomes of functional recovery and sustained health. The semi-supervised association-boosted SNF (abSNF) method will be used to identify subgroups with most significance to executive function performance at baseline. Internal validation of group membership will be performed using bootstrap procedures. Socio-demographic, educational, and life experiential characteristics of each subgroup will be described and placed within the context of existing cross-sectional work.

**Table 4.** Summary of assessment domains and measurements across study projects.

| Project | Domain (instrument/source) | Measures |
| --- | --- | --- |
| 1 | Clinical (MINI, suicidal ideation) + Substance use (TFLB, AUDIT, DUDIT, SDS) | Psychiatric diagnoses, comorbidity, suicidal ideation, childhood adversity, medication, treatment engagement |
|  |  | Recent and lifetime history of (poly)substance use |
|  |  | Age of onset, average yearly use, heaviest period of use |
|  | Cognition (CNS-VS, executive function tasks, other behavioural tasks) | Global cognitive function |
|  |  | Executive function (Inhibition, updating, set shifting) deficits and biases |
|  |  | Risk and reward behaviour |
|  | Functional impairment and quality of life (WHOQOL-BREF, WHODAS) | Physical, psychological, social, and environmental |
|  |  | Cognition, mobility, self-care, getting along, life activities, and participation |
| 2 | Genomics | Assay-based genome-wide genotypes |
|  |  | Assay-based genome-wide methylation profiles |
|  | Plasma biomarkers | Inflammasome proteins (NF-KB, ERK, and p38MAPK, NALP3) |
| | | Pro-inflammatory cytokines (IL-1β, IL-18, and Caspase-1) |
| 3 | Neuroimaging (3T MRI) | Task-based and resting state fMRI |
|  |  | T1-weighted structural MRI |
|  |  | Neuromelanin-sensitive MRI of SN/VTA and LC |
|  |  | Diffusion MRI |
|  |  | Functional near-infrared spectroscopy (for MRI-ineligible) |
| 4 | Intervention (4 week TMS) clinical outcomes | Suicidal ideation (SSI) |
|  |  | Depressive symptoms (HRSD-17) |
|  | Intervention electrophysiological outcomes | Electroencephalogram (EEG)-derived GABA receptor mediated inhibition |
| 6 | General healthcare system utilization (ICES) | Outpatient and inpatient visits, emergency room visits |
|  |  | Maternal health service utilization |
|  |  | Financial costs for hospitalization, procedures, services, drugs, and care |
|  | Addiction and gambling agencies (DATIS) | Demographics, health status, substance use/gambling, treatment intervention, and service utilization |

*Note: This table does not list every measurement from every project and core, though it provides an overview of the types of measurements belonging to each general domain analyzed in P7. P5 is absent, as measures taken from rodent models will not be directly included as features in human sample statistical modeling.*

#### Longitudinal stability of baseline subgroups and subgroup transitions

Within- and across-data type clustering will be performed independently at multiple time points and evaluate the frequencies of individual-level membership transitions. This aim is similar in nature to Aim 2 of Project 1. However, the methods are adapted to account for the increased diversity and dimensionality of input data that will be used to identify latent subgroups. In project 1, latent growth curve modelling will be used to determine if individual trajectories of substance use and functional outcomes are predicted by baseline executive functions. Here in P7, we will extend the foundational work of P1 by examining patient trajectories based on data collected across projects 1-6 and determined using different, network-based methodologies. Critically, we will also identify socio-demographic, educational, and life experiential factors determining inconsistency in group membership over time. Subtypes based on integrative data analyses will be evaluated against more traditional clinical subtypes (such as those from P1) to determine if distributions of key demographics and outcomes are modified by the inclusion of multi-disciplinary input features.

#### Supervised Machine Learning for prognostication

We will apply machine learning methods on features identified by each project across domains to build optimally predictive models of: (1) executive function, (2) social function, (3) suicidality and self-harm, and (4) healthcare system burden (218). Modelling will proceed iteratively, using processes developed by our group (219–221)

## Discussion

There is widespread recognition of the need for integrative and longitudinal research incorporating cognitive, biological, and clinical measures of vulnerability and resilience in SUDs. Within the CDiA Program structure, detailed clinical and cognitive characteristics of a treatment-seeking sample will be integrated with biological markers closely linked to both cognitive dysfunction and SUDs, permitting the identification of critical mechanisms and therapeutic targets. Linkages to healthcare utilization and innovative whole-person modeling approaches will further take this initiative from “neuron-to-neighborhood” and foster integrative and translational research consistent with recent calls to action in this field.

A main strength and innovation of the CDiA Program consists of its translational team and unique collaborative research approach on executive dysfunction for functional recovery and sustained health in outpatients seeking treatment for SUDs (Figure 1). Other conceptual advantages of the Program are the adoption of an individual differences approach, and the recognition that transformation of care can only occur when combining expertise from clinical neuroscience, clinical care, clinical research, and epidemiology/health science research perspectives. As depicted in Figure 1, P1-4 are closely linked investigations with a focus on collecting deep data in complex patients seeking treatment for SUDs; these human subjects research projects emphasize ecological validity and will occur in parallel with P5, which offers the opportunity to investigate associations between EFs, proposed biological underpinnings, and associated pharmacological therapeutics in a highly controlled and efficient design. P6 and P7 extend the generativity of this work, with a focus on healthcare utilization and costs, as well as lived experience perspectives, and the application of highly sophisticated, advanced computational modeling to integrate data across all other projects. To further demonstrate the intended integrated approach across projects, we provide here selected examples of potential for individual projects benefiting and feeding back on strategy, approach and deliverables of other projects (see Table 5).

**Table 5:**
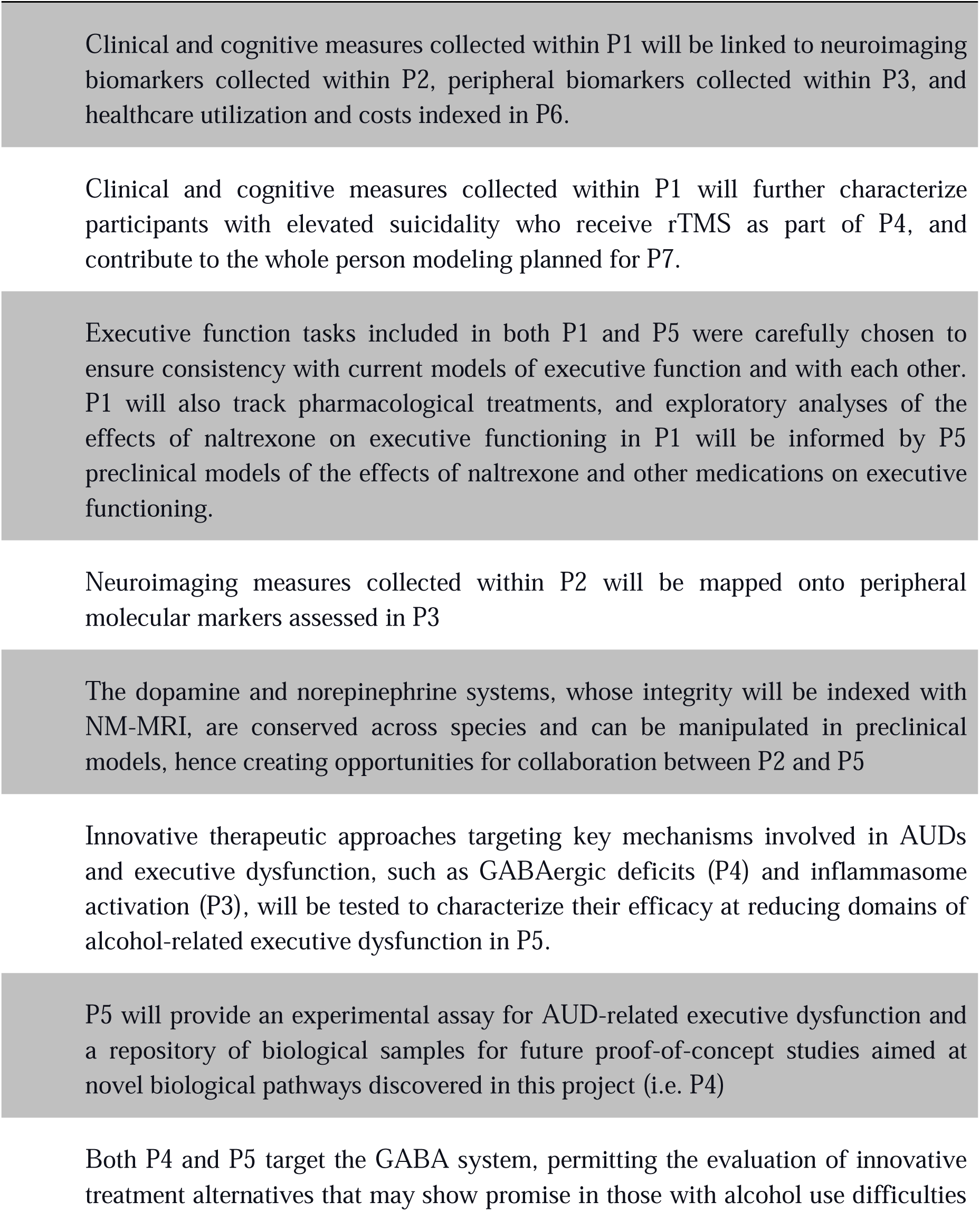
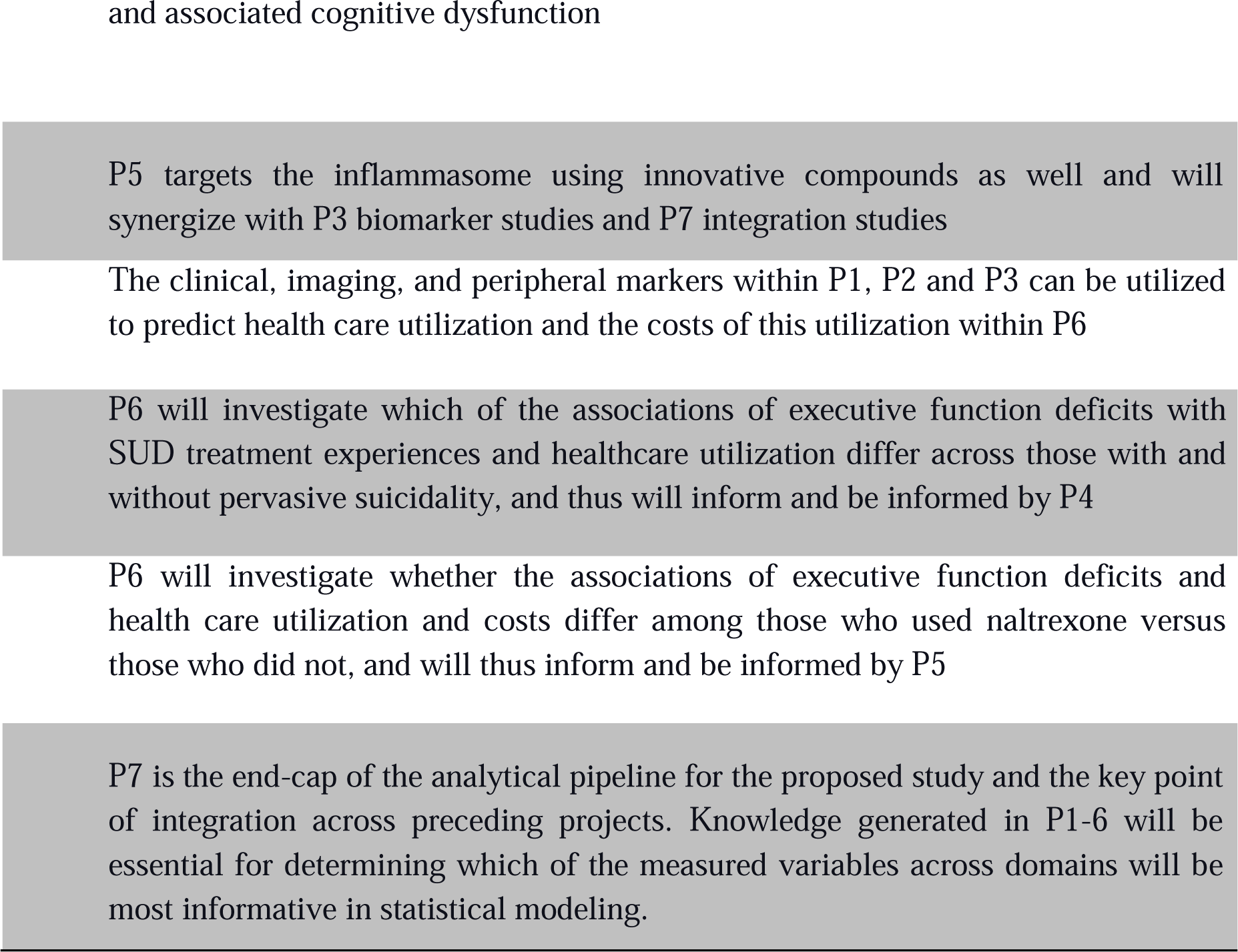
Cross Disciplinary Collaboration Across Projects.

Our inclusive approach to participant recruitment will allow us to recruit a large sample representative of the complex clinical populations typically seen in tertiary care settings. We recognize this will also pose analytical challenges related to sample heterogeneity. Our reliance on multi-level measures of transdiagnostic relevance (e.g., EF, functioning, neural circuit function, inflammatory markers) and our state-of-the-art data integrative approaches outlined in P7 place the CDiA Program in a strong position to harness this heterogeneity to improve understanding of “real-world” presentations of SUDs and their cognitive features. This can in turn accelerate impactful discovery science, clinical translation, and societal/policy changes needed to improve outcomes in this population.

It is anticipated that following the recruitment of this initial cohort, subsequent research will investigate the impact of specific interventions (33,222) on impaired EFs and functional recovery. Thus, it is the vision to use these results to support an “experimental medicine” approach aimed at remediating cognitive dysfunction. Although both psychosocial (e.g., cognitive training) and biological (e.g., pharmacotherapy, noninvasive brain stimulation) interventions are forecasted, the first planned intervention will investigate the efficacy of rTMS on outcomes in SUDs. Peripheral and imaging biomarkers identified in our initial cohort will permit such a nuanced investigation of mediators of clinical outcomes, and in conjunction with preclinical investigations, will inform the development of novel therapeutics (223,224). As an early testament to the generativity of CDiA, two substudies have already been launched: the first aims to assess links between lab-based cognitive and biological measures, and real-world metrics of activity measured by wearable technology, and the second seeks to evaluate the impact of a novel rTMS intervention for treatment of cannabis use disorder (NCT05859347).

Taken together, through its core and embedded projects, the CDiA Program aims to improve precision medicine by understanding the heterogeneity of SUDs, subtyping patients with diverse SUD presentations, discovering underlying neurobiological mechanisms, and identifying targets for novel therapies and therapeutics.

## Ethics Statement

The study was approved by the Research Ethics Board (REB) at the Centre for Addiction and Mental Health (CAMH), REB #: 010/2021. The participants will provide written informed consent to take part in the study. Animal procedures were reviewed and approved by CAMH Animal Care Committee, in accordance with the guidelines of the Canadian Council for Animal Care.

## Authors’ Contributions

PP, LCQ, JDQ,ACR, TDP,YSN, DF, DVC, DV and MES assisted with writing the original draft and review editing. SL assisted with writing the original draft. LQ, KS, WW, DMB, OCM, ANH, VMT, RND SKJ, CR, ES, ASS, BF, PG, JO, ELMV, RNW, KPC and SW assisted with review editing.

DMB, JO, BF, DF, LCQ, DVC, JDF, TDP, ACR, SW, YSN, ES and PP assisted with conceptualization. BF, DF, LCQ, DVC, JDW, TDP, ACR, RND, ELMV worked on investigation. KPC, RNW, PP assisted with software. DF, LCQ, DVC, LQ, RND, RNW, PP assisted with software data curation. DF, LCQ, DVC, JDW, TDP, ACR, SW, YSN, LQ, RND, ELMV, KS, VMT helped with methodology. LCQ, DVC, TDP, ACR provided Supervision. DF, LCQ, DCV, RND assisted with formal analysis. LCQ, ACR, ES, PP, RND, CR helped with project administration. LCQ, DCV, RND, RNW assisted with validation. DF, LCQ, JDW, TDP, ACR, YSN assisted with funding acquisition. DF, LCQ, YSN, RND, ELMV, RNW helps with Resources. DF, LCQ, DCV, YSN, PP, RND, RNW assisted with visualization.

CDiA Program Study Group consists of Lena Quilty, Etienne Sibille, Daniel Felsky, Shannon Lange, Thomas Prevot, Yuliya Nikolova, Anthony Ruocco, Jeffrey Wardell, Erica Vieira, Daphne Voineskos, Isabelle Boileau, Nikki Bozinoff, Daniel Blumberg, Christopher Bowie, Paul Fletcher, Phil Gerretsen, Ahmed Hassan, Colin Hawco, Yarissa Herman, Pamela Kaduri, Paul Kurdyak, Bernard Le Foll, Osnat Melamed, Leanne Quigley, Jürrgen Rehm, Kevin Shield, Matthew Sloan, Victor Tang, Wei Wang and Samantha Wells.

## Data Availability

This is a protocol paper, which does not include any data.

## Acknowledgements

The CDiA Program is supported by a Discovery Fund grant administered through the Centre for Addiction & Mental Health Foundation. The authors gratefully acknowledge the CDiA Program Study Group for their contributions to this research. The authors thank the participants of the CDiA Program. We also thank the CDiA Lived Expertise Research Advisory, CDiA Scientific Advisory Board, and the clinical team members of the CAMH Addictions Program for their contributions and support of this work.

YSN is supported by a Koerner New Scientist Award and a Paul Garfinkel Catalyst Award administered by the CAMH Foundation. DF is supported by CIHR, the Koerner Family Foundation and the Krembil Foundation. DMB receives research support from CIHR, NIMH (R01MH112815;R21MH128815; R01MH1192850), Brain Canada and the Temerty Family through the CAMH Foundation and the Campbell Family Research Institute.

The neurophysiological analysis for Project 5 was partly supported by the Labatt Family Network for Research on the Biology of Depression.

## Footnotes

1 SUD refers to alcohol, cannabis, hallucinogen, opioid, stimulant, and tobacco use disorders.

## References

1. Carvalho AF, Heilig M, Perez A, Probst C, Rehm J. Alcohol use disorders. Lancet (2019) 394:781–792.

2. Rehm J, Shield KD. Global Burden of Disease and the Impact of Mental and Addictive Disorders. Curr Psychiatry Rep (2019) 21:10.

3. Rehm J, Probst C, Falcón LL, Shield KD. “Burden of Disease: The Epidemiological Aspects of Addiction.,” In: el-Guebaly N, Carrà G, Galanter M, Baldacchino AM, editors. Textbook of Addiction Treatment: International Perspectives. Cham: Springer International Publishing (2021). p. 51–64

4. GBD Results. Institute for Health Metrics and Evaluation https://vizhub.healthdata.org/gbd-results/ [Accessed June 6, 2024]

5. Rehm J, Taylor B, Room R. Global burden of disease from alcohol, illicit drugs and tobacco. Drug Alcohol Rev (2006) 25:503–513.

6. Connor JP, Stjepanović D, Le Foll B, Hoch E, Budney AJ, Hall WD. Cannabis use and cannabis use disorder. Nat Rev Dis Primers (2021) 7:16.

7. World Health Organization. Regional Office for the Eastern Mediterranean. Summary report on the regional technical consultation on the working document for development of the action plan (2022-2030) to effectively implement the global strategy to reduce the harmful use of alcohol as a public health priority, virtual meeting, 23 February 2021. World Health Organization. Regional Office for the Eastern Mediterranean (2021). https://apps.who.int/iris/handle/10665/351532 [Accessed March 3, 2023]

8. ARDI alcohol-attributable deaths, US. https://nccd.cdc.gov/DPH_ARDI/Default/Report.aspx?T=AAM&P=612EF325-9B55-442B-AE0C-789B06E3A8D5&R=C877B524-834A-47D5-964D-158FE519C894&M=DB4DAAC0-C9B3-4F92-91A5-A5781DA85B68&F=&D= [Accessed March 20, 2023]

9. Peterson C, Li M, Xu L, Mikosz CA, Luo F. Assessment of Annual Cost of Substance Use Disorder in US Hospitals. JAMA Netw Open (2021) 4:e210242.

10. U.S. Department of Health and Human Services. Substance abuse prevention dollars and cents: A cost-benefit analysis. Barking, England: Lulu.com (2013). 60 p.

11. Hendershot CS, Witkiewitz K, George WH, Marlatt GA. Relapse prevention for addictive behaviors. Subst Abuse Treat Prev Policy (2011) 6:17.

12. Verdejo-Garcia A, Lorenzetti V, Manning V, Piercy H, Bruno R, Hester R, Pennington D, Tolomeo S, Arunogiri S, Bates ME, et al. A Roadmap for Integrating Neuroscience Into Addiction Treatment: A Consensus of the Neuroscience Interest Group of the International Society of Addiction Medicine. Front Psychiatry (2019) 10: doi: 10.3389/fpsyt.2019.00877

13. Kwako LE, Momenan R, Litten RZ, Koob GF, Goldman D. Addictions Neuroclinical Assessment: A Neuroscience-Based Framework for Addictive Disorders. Biol Psychiatry (2016) 80:179–189.

14. Brand M, Wegmann E, Stark R, Müller A, Wölfling K, Robbins TW, Potenza MN. The Interaction of Person-Affect-Cognition-Execution (I-PACE) model for addictive behaviors: Update, generalization to addictive behaviors beyond internet-use disorders, and specification of the process character of addictive behaviors. Neurosci Biobehav Rev (2019) 104:1–10.

15. Banich MT. Executive Function: The Search for an Integrated Account. Curr Dir Psychol Sci (2009) 18:89–94.

16. Jurado MB, Rosselli M. The elusive nature of executive functions: a review of our current understanding. Neuropsychol Rev (2007) 17:213–233.

17. Miller EK, Cohen JD. An integrative theory of prefrontal cortex function. Annu Rev Neurosci (2001) 24:167–202.

18. Miyake A, Friedman NP, Emerson MJ, Witzki AH, Howerter A, Wager TD. The unity and diversity of executive functions and their contributions to complex “Frontal Lobe” tasks: a latent variable analysis. Cogn Psychol (2000) 41:49–100.

19. Hasher L, Lustig C, Zacks R. Inhibitory mechanisms and the control of attention. Variation in working memory (2007) 321:227–249.

20. Nowrangi MA, Lyketsos C, Rao V, Munro CA. Systematic review of neuroimaging correlates of executive functioning: converging evidence from different clinical populations. J Neuropsychiatry Clin Neurosci (2014) 26:114–125.

21. Wise RA, Robble MA. Dopamine and Addiction. Annu Rev Psychol (2020) 71:79–106.

22. Urban NBL, Martinez D. Neurobiology of addiction: insight from neurochemical imaging. Psychiatr Clin North Am (2012) 35:521–541.

23. Ibrahim-Verbaas CA, Bressler J, Debette S, Schuur M, Smith AV, Bis JC, Davies G, Trompet S, Smith JA, Wolf C, et al. GWAS for executive function and processing speed suggests involvement of the CADM2 gene. Mol Psychiatry (2016) 21:189–197.

24. Quigley L, Thiruchselvam T, Quilty LC. Cognitive control biases in depression: A systematic review and meta-analysis. Psychol Bull (2022) 148:662–709.

25. Brorson HH, Ajo Arnevik E, Rand-Hendriksen K, Duckert F. Drop-out from addiction treatment: a systematic review of risk factors. Clin Psychol Rev (2013) 33:1010–1024.

26. Hendershot CS, Wardell JD, Vandervoort J, McPhee MD, Keough MT, Quilty LC. Randomized trial of working memory training as an adjunct to inpatient substance use disorder treatment. Psychol Addict Behav (2018) 32:861–872.

27. Houben K, Wiers RW, Jansen A. Getting a grip on drinking behavior: training working memory to reduce alcohol abuse. Psychol Sci (2011) 22:968–975.

28. Morrison AB, Chein JM. Does working memory training work? The promise and challenges of enhancing cognition by training working memory. Psychon Bull Rev (2011) 18:46–60.

29. Bickel WK, Yi R, Landes RD, Hill PF, Baxter C. Remember the future: working memory training decreases delay discounting among stimulant addicts. Biol Psychiatry (2011) 69:260–265.

30. Dunlop K, Hanlon CA, Downar J. Noninvasive brain stimulation treatments for addiction and major depression. Ann N Y Acad Sci (2017) 1394:31–54.

31. Schluter RS, Daams JG, van Holst RJ, Goudriaan AE. Effects of Non-invasive Neuromodulation on Executive and Other Cognitive Functions in Addictive Disorders: A Systematic Review. Front Neurosci (2018) 12:642.

32. Stein ER, Gibson BC, Votaw VR, Wilson AD, Clark VP, Witkiewitz K. Non-invasive brain stimulation in substance use disorders: implications for dissemination to clinical settings. Curr Opin Psychol (2019) 30:6–10.

33. Lechner WV, Sidhu NK, Kittaneh AA, Anand A. Interventions with potential to target executive function deficits in addiction: current state of the literature. Curr Opin Psychol (2019) 30:24–28.

34. Verdejo-Garcia A, Garcia-Fernandez G, Dom G. Cognition and addiction. Dialogues Clin Neurosci (2019) 21:281–290.

35. Kim-Spoon J, Kahn RE, Lauharatanahirun N, Deater-Deckard K, Bickel WK, Chiu PH, King-Casas B. Executive functioning and substance use in adolescence: Neurobiological and behavioral perspectives. Neuropsychologia (2017) 100:79–92.

36. Kuhns L, Kroon E, Lesscher H, Mies G, Cousijn J. Age-related differences in the effect of chronic alcohol on cognition and the brain: a systematic review. Transl Psychiatry (2022) 12:345.

37. Teichner G, Horner MD, Roitzsch JC, Herron J, Thevos A. Substance abuse treatment outcomes for cognitively impaired and intact outpatients. Addict Behav (2002) 27:751–763.

38. Mojtabai R, Olfson M, Mechanic D. Perceived need and help-seeking in adults with mood, anxiety, or substance use disorders. Arch Gen Psychiatry (2002) 59:77–84.

39. Graham K, Cheng J, Bernards S, Wells S, Rehm J, Kurdyak P. How Much Do Mental Health and Substance Use/Addiction Affect Use of General Medical Services? Extent of Use, Reason for Use, and Associated Costs. Can J Psychiatry (2017) 62:48–56.

40. Moustafa AA, Morris AN, Nandrino JL, Misiak B, Szewczuk-Bogusławska M, Frydecka D, El Haj M. Not all drugs are created equal: impaired future thinking in opiate, but not alcohol, users. Exp Brain Res (2018) 236:2971–2981.

41. Rolland B, D’Hondt F, Montègue S, Brion M, Peyron E, D’Aviau de Ternay J, de Timary P, Nourredine M, Maurage P. A Patient-Tailored Evidence-Based Approach for Developing Early Neuropsychological Training Programs in Addiction Settings. Neuropsychol Rev (2019) 29:103–115.

42. Quilty LC. Commentary on Verdejo-Garcia, et al.: Cognitive training and remediation: identifying priorities for intervention research. Addiction (2023) 118:952–953.

43. Lindgren KP, Hendershot CS, Ramirez JJ, Bernat E, Rangel-Gomez M, Peterson KP, Murphy JG. A dual process perspective on advances in cognitive science and alcohol use disorder. Clin Psychol Rev (2019) 69:83–96.

44. Wiers RW, Gladwin TE, Hofmann W, Salemink E, Ridderinkhof KR. Cognitive Bias Modification and Cognitive Control Training in Addiction and Related Psychopathology: Mechanisms, Clinical Perspectives, and Ways Forward. Clin Psychol Sci (2013) 1:192–212.

45. Gould TJ. Addiction and cognition. Addict Sci Clin Pract (2010) 5:4–14.

46. Kwako LE, Bickel WK, Goldman D. Addiction Biomarkers: Dimensional Approaches to Understanding Addiction. Trends Mol Med (2018) 24:121–128.

47. Kwako LE, Momenan R, Grodin EN, Litten RZ, Koob GF, Goldman D. Addictions Neuroclinical Assessment: A reverse translational approach. Neuropharmacology (2017) 122:254–264.

48. Domínguez-Salas S, Díaz-Batanero C, Lozano-Rojas OM, Verdejo-García A. Impact of general cognition and executive function deficits on addiction treatment outcomes: Systematic review and discussion of neurocognitive pathways. Neurosci Biobehav Rev (2016) 71:772–801.

49. McCabe RE, Milosevic I, Rowa K, Shnaider P, Pawluk EJ, Antony MM. Diagnostic Assessment Research Tool. Psychol Assess doi: 10.1037/t81500-000

50. Sobell LC, Ontario. Addiction Research Foundation, Sobell MB. Timeline Followback: User’s Guide. Addiction Research Foundation = Fondation de la recherche sur la toxicomanie (1996). 109 p.

51. Gualtieri CT, Johnson LG. Reliability and validity of a computerized neurocognitive test battery, CNS Vital Signs. Arch Clin Neuropsychol (2006) 21:623–643.

52. Bechara A, Damasio AR, Damasio H, Anderson SW. Insensitivity to future consequences following damage to human prefrontal cortex. Cognition (1994) 50:7–15.

53. Myerson J, Green L, Scott Hanson J, Holt DD, Estle SJ. Discounting delayed and probabilistic rewards: Processes and traits. J Econ Psychol (2003) 24:619–635.

54. Myerson J, Green L, Morris J. Modeling the effect of reward amount on probability discounting. J Exp Anal Behav (2011) 95:175–187.

55. MacKillop J, Weafer J, C Gray J, Oshri A, Palmer A, de Wit H. The latent structure of impulsivity: impulsive choice, impulsive action, and impulsive personality traits. Psychopharmacology (2016) 233:3361–3370.

56. Lundqvist D, Flykt A, Öhman A. Karolinska Directed Emotional Faces. Cognition and Emotion doi: 10.1037/t27732-000

57. Eriksen BA, Eriksen CW. Effects of noise letters upon the identification of a target letter in a nonsearch task. Percept Psychophys (1974) 16:143–149.

58. Fenske MJ, Eastwood JD. Modulation of focused attention by faces expressing emotion: evidence from flanker tasks. Emotion (2003) 3:327–343.

59. Zetsche U, D’Avanzato C, Joormann J. Depression and rumination: relation to components of inhibition. Cogn Emot (2012) 26:758–767.

60. Zetsche U, Joormann J. Components of interference control predict depressive symptoms and rumination cross-sectionally and at six months follow-up. J Behav Ther Exp Psychiatry (2011) 42:65–73.

61. Levens SM, Gotlib IH. Updating positive and negative stimuli in working memory in depression. J Exp Psychol Gen (2010) 139:654–664.

62. Pe ML, Raes F, Kuppens P. The cognitive building blocks of emotion regulation: ability to update working memory moderates the efficacy of rumination and reappraisal on emotion. PLoS One (2013) 8:e69071.

63. Morris N, Jones DM. Memory updating in working memory: The role of the central executive. Br J Psychol (1990) 81:111–121.

64. Reineberg AE, Andrews-Hanna JR, Depue BE, Friedman NP, Banich MT. Resting-state networks predict individual differences in common and specific aspects of executive function. Neuroimage (2015) 104:69–78.

65. Reineberg AE, Banich MT. Functional connectivity at rest is sensitive to individual differences in executive function: A network analysis. Hum Brain Mapp (2016) 37:2959– 2975.

66. Rieck JR, Baracchini G, Grady CL. Contributions of Brain Function and Structure to Three Different Domains of Cognitive Control in Normal Aging. J Cogn Neurosci (2021) 33:1811–1832.

67. He L, Zhuang K, Chen Q, Wei D, Chen X, Fan J, Qiu J. Unity and diversity of neural representation in executive functions. J Exp Psychol Gen (2021) 150:2193–2207.

68. Braver TS. The variable nature of cognitive control: a dual mechanisms framework. Trends Cogn Sci (2012) 16:106–113.

69. Sakai K. Task set and prefrontal cortex. Annu Rev Neurosci (2008) 31:219–245.

70. Dosenbach NUF, Fair DA, Cohen AL, Schlaggar BL, Petersen SE. A dual-networks architecture of top-down control. Trends Cogn Sci (2008) 12:99–105.

71. Zhang R, Geng X, Lee TMC. Large-scale functional neural network correlates of response inhibition: an fMRI meta-analysis. Brain Struct Funct (2017) 222:3973–3990.

72. Derrfuss J, Brass M, Neumann J, von Cramon DY. Involvement of the inferior frontal junction in cognitive control: meta-analyses of switching and Stroop studies. Hum Brain Mapp (2005) 25:22–34.

73. Wager TD, Jonides J, Reading S. Neuroimaging studies of shifting attention: a meta-analysis. Neuroimage (2004) 22:1679–1693.

74. O’Reilly RC, Frank MJ. Making working memory work: a computational model of learning in the prefrontal cortex and basal ganglia. Neural Comput (2006) 18:283–328.

75. McNab F, Klingberg T. Prefrontal cortex and basal ganglia control access to working memory. Nat Neurosci (2008) 11:103–107.

76. Emch M, von Bastian CC, Koch K. Neural Correlates of Verbal Working Memory: An fMRI Meta-Analysis. Front Hum Neurosci (2019) 13:180.

77. Bettcher BM, Mungas D, Patel N, Elofson J, Dutt S, Wynn M, Watson CL, Stephens M, Walsh CM, Kramer JH. Neuroanatomical substrates of executive functions: Beyond prefrontal structures. Neuropsychologia (2016) 85:100–109.

78. Yuan P, Raz N. Prefrontal cortex and executive functions in healthy adults: a meta-analysis of structural neuroimaging studies. Neurosci Biobehav Rev (2014) 42:180–192.

79. Klugah-Brown B, Di X, Zweerings J, Mathiak K, Becker B, Biswal B. Common and separable neural alterations in substance use disorders: A coordinate-based meta-analyses of functional neuroimaging studies in humans. Hum Brain Mapp (2020) 41:4459–4477.

80. Navarri X, Afzali MH, Lavoie J, Sinha R, Stein DJ, Momenan R, Veltman DJ, Korucuoglu O, Sjoerds Z, van Holst RJ, et al. How do substance use disorders compare to other psychiatric conditions on structural brain abnormalities? A cross-disorder meta-analytic comparison using the ENIGMA consortium findings. Hum Brain Mapp (2022) 43:399–413.

81. Yan H, Xiao S, Fu S, Gong J, Qi Z, Chen G, Chen P, Tang G, Su T, Yang Z, et al. Functional and structural brain abnormalities in substance use disorder: A multimodal meta-analysis of neuroimaging studies. Acta Psychiatr Scand (2023) 147:345–359.

82. Dugré JR, Orban P, Potvin S. Disrupted functional connectivity of the brain reward system in substance use problems: A meta-analysis of functional neuroimaging studies. Addict Biol (2023) 28:e13257.

83. Casey BJ, Cannonier T, Conley MI, Cohen AO, Barch DM, Heitzeg MM, Soules ME, Teslovich T, Dellarco DV, Garavan H, et al. The Adolescent Brain Cognitive Development (ABCD) study: Imaging acquisition across 21 sites. Dev Cogn Neurosci (2018) 32:43–54.

84. Dickie EW, Ameis SH, Boileau I, Diaconescu AO, Felsky D, Goldstein BI, Goncalves V, Griffiths JD, Haltigan JD, Husain MO, et al. Neuroimaging & Biosample Collection in the Toronto Adolescent and Youth (TAY) Cohort Study: Rationale, Methods, and Early Data. Biol Psychiatry Cogn Neurosci Neuroimaging (2023) doi: 10.1016/j.bpsc.2023.10.013

85. Agarwal SM, Dissanayake J, Agid O, Bowie C, Brierley N, Chintoh A, De Luca V, Diaconescu A, Gerretsen P, Graff-Guerrero A, et al. Characterization and prediction of individual functional outcome trajectories in schizophrenia spectrum disorders (PREDICTS study): Study protocol. PLoS One (2023) 18:e0288354.

86. Cassidy CM, Carpenter KM, Konova AB, Cheung V, Grassetti A, Zecca L, Abi-Dargham A, Martinez D, Horga G. Evidence for Dopamine Abnormalities in the Substantia Nigra in Cocaine Addiction Revealed by Neuromelanin-Sensitive MRI. Am J Psychiatry (2020) 177:1038–1047.

87. Cassidy CM, Zucca FA, Girgis RR, Baker SC, Weinstein JJ, Sharp ME, Bellei C, Valmadre A, Vanegas N, Kegeles LS, et al. Neuromelanin-sensitive MRI as a noninvasive proxy measure of dopamine function in the human brain. Proc Natl Acad Sci U S A (2019) 116:5108–5117.

88. Calarco N, Cassidy CM, Selby B, Hawco C, Voineskos AN, Diniz BS, Nikolova YS. Associations between locus coeruleus integrity and diagnosis, age, and cognitive performance in older adults with and without late-life depression: An exploratory study. Neuroimage Clin (2022) 36:103182.

89. Foster SL, Weinshenker D. “Chapter 15 - The Role of Norepinephrine in Drug Addiction: Past, Present, and Future.,” In: Torregrossa M, editor. Neural Mechanisms of Addiction. Academic Press (2019). p. 221–236

90. Chen Z, Lei X, Ding C, Li H, Chen A. The neural mechanisms of semantic and response conflicts: an fMRI study of practice-related effects in the Stroop task. Neuroimage (2013) 66:577–584.

91. Weissman DH, Woldorff MG, Hazlett CJ, Mangun GR. Effects of practice on executive control investigated with fMRI. Brain Res Cogn Brain Res (2002) 15:47–60.

92. Turner BO, Paul EJ, Miller MB, Barbey AK. Small sample sizes reduce the replicability of task-based fMRI studies. Commun Biol (2018) 1:62.

93. Copersino ML. Cognitive Mechanisms and Therapeutic Targets of Addiction. Curr Opin Behav Sci (2017) 13:91–98.

94. Bough KJ, Pollock JD. Defining Substance Use Disorders: The Need for Peripheral Biomarkers. Trends Mol Med (2018) 24:109–120.

95. Torkamani A, Wineinger NE, Topol EJ. The personal and clinical utility of polygenic risk scores. Nat Rev Genet (2018) 19:581–590.

96. Khera AV, Chaffin M, Aragam KG, Haas ME, Roselli C, Choi SH, Natarajan P, Lander ES, Lubitz SA, Ellinor PT, et al. Genome-wide polygenic scores for common diseases identify individuals with risk equivalent to monogenic mutations. Nat Genet (2018) 50:1219–1224.

97. Redwine LS, Pung MA, Wilson K, Chinh K, Duffy AR. Differential Peripheral Inflammatory Factors Associated with Cognitive Function in Patients with Heart Failure. Neuroimmunomodulation (2018) 25:146–152.

98. Ersche KD, Roiser JP, Lucas M, Domenici E, Robbins TW, Bullmore ET. Peripheral biomarkers of cognitive response to dopamine receptor agonist treatment. Psychopharmacology (2011) 214:779–789.

99. Crews FT, Zou J, Qin L. Induction of innate immune genes in brain create the neurobiology of addiction. Brain Behav Immun (2011) 25 Suppl 1:S4–S12.

100. Umhau JC, Schwandt M, Solomon MG, Yuan P, Nugent A, Zarate CA, Drevets WC, Hall SD, George DT, Heilig M. Cerebrospinal fluid monocyte chemoattractant protein-1 in alcoholics: support for a neuroinflammatory model of chronic alcoholism. Alcohol Clin Exp Res (2014) 38:1301–1306.

101. Portelli J, Wiers CE, Li X, Deschaine SL, McDiarmid GR, Bermpohl F, Leggio L. Peripheral proinflammatory markers are upregulated in abstinent alcohol-dependent patients but are not affected by cognitive bias modification: Preliminary findings. Drug Alcohol Depend (2019) 204:107553.

102. Girard M, Malauzat D, Nubukpo P. Serum inflammatory molecules and markers of neuronal damage in alcohol-dependent subjects after withdrawal. World J Biol Psychiatry (2019) 20:76–90.

103. Zaparte A, Schuch JB, Viola TW, Baptista TAS, Beidacki AS, do Prado CH, Sanvicente-Vieira B, Bauer ME, Grassi-Oliveira R. Cocaine Use Disorder Is Associated With Changes in Th1/Th2/Th17 Cytokines and Lymphocytes Subsets. Front Immunol (2019) 10:2435.

104. Levandowski ML, Hess ARB, Grassi-Oliveira R, de Almeida RMM. Plasma interleukin-6 and executive function in crack cocaine-dependent women. Neurosci Lett (2016) 628:85– 90.

105. Wang T-Y, Lee S-Y, Chang Y-H, Chen S-L, Chen PS, Chu C-H, Huang S-Y, Tzeng N-S, Lee IH, Chen KC, et al. Correlation of cytokines, BDNF levels, and memory function in patients with opioid use disorder undergoing methadone maintenance treatment. Drug Alcohol Depend (2018) 191:6–13.

106. Lu R-B, Wang T-Y, Lee S-Y, Chen S-L, Chang Y-H, See Chen P, Lin S-H, Chu C-H, Huang S-Y, Tzeng N-S, et al. Correlation between interleukin-6 levels and methadone maintenance therapy outcomes. Drug Alcohol Depend (2019) 204:107516.

107. Bayazit H, Selek S, Karababa IF, Cicek E, Aksoy N. Evaluation of Oxidant/Antioxidant Status and Cytokine Levels in Patients with Cannabis Use Disorder. Clin Psychopharmacol Neurosci (2017) 15:237–242.

108. Liu L, Chan C. The role of inflammasome in Alzheimer’s disease. Ageing Res Rev (2014) 15:6–15.

109. Minkiewicz J, de Rivero Vaccari JP, Keane RW. Human astrocytes express a novel NLRP2 inflammasome. Glia (2013) 61:1113–1121.

110. Silverman WR, de Rivero Vaccari JP, Locovei S, Qiu F, Carlsson SK, Scemes E, Keane RW, Dahl G. The pannexin 1 channel activates the inflammasome in neurons and astrocytes. J Biol Chem (2009) 284:18143–18151.

111. Halle A, Hornung V, Petzold GC, Stewart CR, Monks BG, Reinheckel T, Fitzgerald KA, Latz E, Moore KJ, Golenbock DT. The NALP3 inflammasome is involved in the innate immune response to amyloid-beta. Nat Immunol (2008) 9:857–865.

112. Ye T, Meng X, Wang R, Zhang C, He S, Sun G, Sun X. Gastrodin Alleviates Cognitive Dysfunction and Depressive-Like Behaviors by Inhibiting ER Stress and NLRP3 Inflammasome Activation in db/db Mice. Int J Mol Sci (2018) 19: doi: 10.3390/ijms19123977

113. Zhuang J, Wen X, Zhang Y-Q, Shan Q, Zhang Z-F, Zheng G-H, Fan S-H, Li M-Q, Wu D-M, Hu B, et al. TDP-43 upregulation mediated by the NLRP3 inflammasome induces cognitive impairment in 2 2’,4,4’-tetrabromodiphenyl ether (BDE-47)-treated mice. Brain Behav Immun (2017) 65:99–110.

114. Forlenza OV, Diniz BS, Talib LL, Mendonça VA, Ojopi EB, Gattaz WF, Teixeira AL. Increased serum IL-1beta level in Alzheimer’s disease and mild cognitive impairment. Dement Geriatr Cogn Disord (2009) 28:507–512.

115. Diniz BS, Teixeira AL, Talib L, Gattaz WF, Forlenza OV. Interleukin-1beta serum levels is increased in antidepressant-free elderly depressed patients. Am J Geriatr Psychiatry (2010) 18:172–176.

116. Karagüzel EÖ, Arslan FC, Uysal EK, Demir S, Aykut DS, Tat M, Karahan SC. Blood levels of interleukin-1 beta, interleukin-6 and tumor necrosis factor-alpha and cognitive functions in patients with obsessive compulsive disorder. Compr Psychiatry (2019) 89:61– 66.

117. Tian S, Huang R, Han J, Cai R, Guo D, Lin H, Wang J, Wang S. Increased plasma Interleukin-1β level is associated with memory deficits in type 2 diabetic patients with mild cognitive impairment. Psychoneuroendocrinology (2018) 96:148–154.

118. O’Reardon JP, Solvason HB, Janicak PG, Sampson S, Isenberg KE, Nahas Z, McDonald WM, Avery D, Fitzgerald PB, Loo C, et al. Efficacy and safety of transcranial magnetic stimulation in the acute treatment of major depression: a multisite randomized controlled trial. Biol Psychiatry (2007) 62:1208–1216.

119. Carpentier PJ, Krabbe PFM, van Gogh MT, Knapen LJM, Buitelaar JK, de Jong CAJ. Psychiatric comorbidity reduces quality of life in chronic methadone maintained patients. Am J Addict (2009) 18:470–480.

120. Darke S, Ross J. Suicide among heroin users: rates, risk factors and methods. Addiction (2002) 97:1383–1394.

121. Rounsaville BJ, Weissman MM, Crits-Christoph K, Wilber C, Kleber H. Diagnosis and symptoms of depression in opiate addicts. Course and relationship to treatment outcome. Arch Gen Psychiatry (1982) 39:151–156.

122. Rounsaville BJ, Weissman MM, Kleber H, Wilber C. Heterogeneity of psychiatric diagnosis in treated opiate addicts. Arch Gen Psychiatry (1982) 39:161–168.

123. Warner-Smith M, Darke S, Day C. Morbidity associated with non-fatal heroin overdose. Addiction (2002) 97:963–967.

124. Marzuk PM, Hartwell N, Leon AC, Portera L. Executive functioning in depressed patients with suicidal ideation. Acta Psychiatr Scand (2005) 112:294–301.

125. Koenigs M, Grafman J. The functional neuroanatomy of depression: distinct roles for ventromedial and dorsolateral prefrontal cortex. Behav Brain Res (2009) 201:239–243.

126. Spagnolo PA, Goldman D. Neuromodulation interventions for addictive disorders: challenges, promise, and roadmap for future research. Brain (2017) 140:1183–1203.

127. Volkow ND, Koob GF, Croyle RT, Bianchi DW, Gordon JA, Koroshetz WJ, Pérez-Stable EJ, Riley WT, Bloch MH, Conway K, et al. The conception of the ABCD study: From substance use to a broad NIH collaboration. Dev Cogn Neurosci (2018) 32:4–7.

128. van ’t Wout M, Kahn RS, Sanfey AG, Aleman A. Repetitive transcranial magnetic stimulation over the right dorsolateral prefrontal cortex affects strategic decision-making. Neuroreport (2005) 16:1849–1852.

129. George MS, Lisanby SH, Avery D, McDonald WM, Durkalski V, Pavlicova M, Anderson B, Nahas Z, Bulow P, Zarkowski P, et al. Daily left prefrontal transcranial magnetic stimulation therapy for major depressive disorder: a sham-controlled randomized trial. Arch Gen Psychiatry (2010) 67:507–516.

130. O’Reardon JP, Solvason HB, Janicak PG, Sampson S, Isenberg KE, Nahas Z, McDonald WM, Avery D, Fitzgerald PB, Loo C, et al. Reply regarding “efficacy and safety of transcranial magnetic stimulation in the acute treatment of major depression: a multisite randomized controlled trial.” Biol Psychiatry (2010) 67:e15–7.

131. Li C-T, Chen M-H, Juan C-H, Huang H-H, Chen L-F, Hsieh J-C, Tu P-C, Bai Y-M, Tsai S-J, Lee Y-C, et al. Efficacy of prefrontal theta-burst stimulation in refractory depression: a randomized sham-controlled study. Brain (2014) 137:2088–2098.

132. Huang Y-Z, Edwards MJ, Rounis E, Bhatia KP, Rothwell JC. Theta burst stimulation of the human motor cortex. Neuron (2005) 45:201–206.

133. Uncapher H, Gallagher-Thompson D, Osgood NJ, Bongar B. Hopelessness and suicidal ideation in older adults. Gerontologist (1998) 38:62–70.

134. Papakostas GI, Petersen T, Pava J, Masson E, Worthington JJ 3rd, Alpert JE, Fava M, Nierenberg AA. Hopelessness and suicidal ideation in outpatients with treatment-resistant depression: prevalence and impact on treatment outcome. J Nerv Ment Dis (2003) 191:444– 449.

135. Insel T, Cuthbert B, Garvey M, Heinssen R, Pine DS, Quinn K, Sanislow C, Wang P. Research domain criteria (RDoC): toward a new classification framework for research on mental disorders. Am J Psychiatry (2010) 167:748–751.

136. Murphy SC, Palmer LM, Nyffeler T, Müri RM, Larkum ME. Transcranial magnetic stimulation (TMS) inhibits cortical dendrites. Elife (2016) 5: doi: 10.7554/eLife.13598

137. Bender S, Basseler K, Sebastian I, Resch F, Kammer T, Oelkers-Ax R, Weisbrod M. Electroencephalographic response to transcranial magnetic stimulation in children: Evidence for giant inhibitory potentials. Ann Neurol (2005) 58:58–67.

138. Daskalakis ZJ, Farzan F, Barr MS, Maller JJ, Chen R, Fitzgerald PB. Long-interval cortical inhibition from the dorsolateral prefrontal cortex: a TMS-EEG study. Neuropsychopharmacology (2008) 33:2860–2869.

139. Farzan F, Barr MS, Hoppenbrouwers SS, Fitzgerald PB, Chen R, Pascual-Leone A, Daskalakis ZJ. The EEG correlates of the TMS-induced EMG silent period in humans. Neuroimage (2013) 83:120–134.

140. Nikulin VV, Kicić D, Kähkönen S, Ilmoniemi RJ. Modulation of electroencephalographic responses to transcranial magnetic stimulation: evidence for changes in cortical excitability related to movement. Eur J Neurosci (2003) 18:1206–1212.

141. Premoli I, Castellanos N, Rivolta D, Belardinelli P, Bajo R, Zipser C, Espenhahn S, Heidegger T, Müller-Dahlhaus F, Ziemann U. TMS-EEG signatures of GABAergic neurotransmission in the human cortex. J Neurosci (2014) 34:5603–5612.

142. Daskalakis ZJ, Möller B, Christensen BK, Fitzgerald PB, Gunraj C, Chen R. The effects of repetitive transcranial magnetic stimulation on cortical inhibition in healthy human subjects. Exp Brain Res (2006) 174:403–412.

143. Chung SW, Lewis BP, Rogasch NC, Saeki T, Thomson RH, Hoy KE, Bailey NW, Fitzgerald PB. Demonstration of short-term plasticity in the dorsolateral prefrontal cortex with theta burst stimulation: A TMS-EEG study. Clin Neurophysiol (2017) 128:1117–1126.

144. Chung SW, Rogasch NC, Hoy KE, Sullivan CM, Cash RFH, Fitzgerald PB. Impact of different intensities of intermittent theta burst stimulation on the cortical properties during TMS-EEG and working memory performance. Hum Brain Mapp (2018) 39:783–802.

145. Voineskos D, Blumberger DM, Zomorrodi R, Rogasch NC, Farzan F, Foussias G, Rajji TK, Daskalakis ZJ. Altered Transcranial Magnetic Stimulation-Electroencephalographic Markers of Inhibition and Excitation in the Dorsolateral Prefrontal Cortex in Major Depressive Disorder. Biol Psychiatry (2019) 85:477–486.

146. Brooks H, Goodman MS, Bowie CR, Zomorrodi R, Blumberger DM, Butters MA, Daskalakis ZJ, Fischer CE, Flint A, Herrmann N, et al. Theta-gamma coupling and ordering information: a stable brain-behavior relationship across cognitive tasks and clinical conditions. Neuropsychopharmacology (2020) 45:2038–2047.

147. Noda Y, Zomorrodi R, Saeki T, Rajji TK, Blumberger DM, Daskalakis ZJ, Nakamura M. Resting-state EEG gamma power and theta-gamma coupling enhancement following high-frequency left dorsolateral prefrontal rTMS in patients with depression. Clin Neurophysiol (2017) 128:424–432.

148. Blumberger DM, Vila-Rodriguez F, Thorpe KE, Feffer K, Noda Y, Giacobbe P, Knyahnytska Y, Kennedy SH, Lam RW, Daskalakis ZJ, et al. Effectiveness of theta burst versus high-frequency repetitive transcranial magnetic stimulation in patients with depression (THREE-D): a randomised non-inferiority trial. Lancet (2018) 391:1683–1692.

149. Bakker N, Shahab S, Giacobbe P, Blumberger DM, Daskalakis ZJ, Kennedy SH, Downar J. rTMS of the dorsomedial prefrontal cortex for major depression: safety, tolerability, effectiveness, and outcome predictors for 10 Hz versus intermittent theta-burst stimulation. Brain Stimul (2015) 8:208–215.

150. Dunlop K, Gaprielian P, Blumberger D, Daskalakis ZJ, Kennedy SH, Giacobbe P, Downar J. MRI-guided dmPFC-rTMS as a Treatment for Treatment-resistant Major Depressive Disorder. J Vis Exp (2015)e53129.

151. Keel JC, Smith MJ, Wassermann EM. A safety screening questionnaire for transcranial magnetic stimulation. Clin Neurophysiol (2001) 112:720.

152. Kaster TS, Daskalakis ZJ, Noda Y, Knyahnytska Y, Downar J, Rajji TK, Levkovitz Y, Zangen A, Butters MA, Mulsant BH, et al. Efficacy, tolerability, and cognitive effects of deep transcranial magnetic stimulation for late-life depression: a prospective randomized controlled trial. Neuropsychopharmacology (2018) 43:2231–2238.

153. Hamilton M. Development of a rating scale for primary depressive illness. Br J Soc Clin Psychol (1967) 6:278–296.

154. Arias SA, Miller I, Camargo CA Jr, Sullivan AF, Goldstein AB, Allen MH, Manton AP, Boudreaux ED. Factors Associated With Suicide Outcomes 12 Months After Screening Positive for Suicide Risk in the Emergency Department. Psychiatr Serv (2016) 67:206–213.

155. Ionescu DF, Swee MB, Pavone KJ, Taylor N, Akeju O, Baer L, Nyer M, Cassano P, Mischoulon D, Alpert JE, et al. Rapid and Sustained Reductions in Current Suicidal Ideation Following Repeated Doses of Intravenous Ketamine: Secondary Analysis of an Open-Label Study. J Clin Psychiatry (2016) 77:e719–25.

156. Harvey PD, Posner K, Rajeevan N, Yershova KV, Aslan M, Concato J. Suicidal ideation and behavior in US veterans with schizophrenia or bipolar disorder. J Psychiatr Res (2018) 102:216–222.

157. Legarreta M, Graham J, North L, Bueler CE, McGlade E, Yurgelun-Todd D. DSM-5 posttraumatic stress disorder symptoms associated with suicide behaviors in veterans. Psychol Trauma (2015) 7:277–285.

158. Gwaltney C, Mundt JC, Greist JH, Paty J, Tiplady B. Interactive Voice Response and Text-based Self-report Versions of the Electronic Columbia-Suicide Severity Rating Scale Are Equivalent. Innov Clin Neurosci (2017) 14:17–23.

159. Hesdorffer DC, French JA, Posner K, DiVentura B, Pollard JR, Sperling MR, Harden CL, Krauss GL, Kanner AM. Suicidal ideation and behavior screening in intractable focal epilepsy eligible for drug trials. Epilepsia (2013) 54:879–887.

160. Kerr DCR, DeGarmo DS, Leve LD, Chamberlain P. Juvenile justice girls’ depressive symptoms and suicidal ideation 9 years after Multidimensional Treatment Foster Care. J Consult Clin Psychol (2014) 82:684–693.

161. Kerr DCR, Gibson B, Leve LD, Degarmo DS. Young adult follow-up of adolescent girls in juvenile justice using the Columbia suicide severity rating scale. Suicide Life Threat Behav (2014) 44:113–129.

162. Sun Y, Farzan F, Mulsant BH, Rajji TK, Fitzgerald PB, Barr MS, Downar J, Wong W, Blumberger DM, Daskalakis ZJ. Indicators for Remission of Suicidal Ideation Following Magnetic Seizure Therapy in Patients With Treatment-Resistant Depression. JAMA Psychiatry (2016) 73:337–345.

163. Farzan F, Barr MS, Levinson AJ, Chen R, Wong W, Fitzgerald PB, Daskalakis ZJ. Reliability of long-interval cortical inhibition in healthy human subjects: a TMS-EEG study. J Neurophysiol (2010) 104:1339–1346.

164. Lioumis P, Kicić D, Savolainen P, Mäkelä JP, Kähkönen S. Reproducibility of TMS-Evoked EEG responses. Hum Brain Mapp (2009) 30:1387–1396.

165. Goodman MS, Kumar S, Zomorrodi R, Ghazala Z, Cheam ASM, Barr MS, Daskalakis ZJ, Blumberger DM, Fischer C, Flint A, et al. Theta-Gamma Coupling and Working Memory in Alzheimer’s Dementia and Mild Cognitive Impairment. Front Aging Neurosci (2018) 10:101.

166. Charlton AJ, May C, Luikinga SJ, Burrows EL, Hyun Kim J, Lawrence AJ, Perry CJ. Chronic voluntary alcohol consumption causes persistent cognitive deficits and cortical cell loss in a rodent model. Sci Rep (2019) 9:18651.

167. Broadwater M, Varlinskaya EI, Spear LP. Chronic intermittent ethanol exposure in early adolescent and adult male rats: effects on tolerance, social behavior, and ethanol intake. Alcohol Clin Exp Res (2011) 35:1392–1403.

168. Prévot T, Sibille E. Altered GABA-mediated information processing and cognitive dysfunctions in depression and other brain disorders. Mol Psychiatry (2021) 26:151–167.

169. Lang Y, Chu F, Shen D, Zhang W, Zheng C, Zhu J, Cui L. Role of Inflammasomes in Neuroimmune and Neurodegenerative Diseases: A Systematic Review. Mediators Inflamm (2018) 2018:1549549.

170. Lippai D, Bala S, Petrasek J, Csak T, Levin I, Kurt-Jones EA, Szabo G. Alcohol-induced IL-1β in the brain is mediated by NLRP3/ASC inflammasome activation that amplifies neuroinflammation. J Leukoc Biol (2013) 94:171–182.

171. Fitzgerald PJ. Elevated Norepinephrine may be a Unifying Etiological Factor in the Abuse of a Broad Range of Substances: Alcohol, Nicotine, Marijuana, Heroin, Cocaine, and Caffeine. Subst Abuse (2013) 7:171–183.

172. Fredriksson I, Jayaram-Lindström N, Wirf M, Nylander E, Nyström E, Jardemark K, Steensland P. Evaluation of guanfacine as a potential medication for alcohol use disorder in long-term drinking rats: behavioral and electrophysiological findings. Neuropsychopharmacology (2015) 40:1130–1140.

173. Newcorn JH, Harpin V, Huss M, Lyne A, Sikirica V, Johnson M, Ramos-Quiroga JA, van Stralen J, Dutray B, Sreckovic S, et al. Extended-release guanfacine hydrochloride in 6-17-year olds with ADHD: a randomised-withdrawal maintenance of efficacy study. J Child Psychol Psychiatry (2016) 57:717–728.

174. Setiawan E, Pihl RO, Cox SML, Gianoulakis C, Palmour RM, Benkelfat C, Leyton M. The effect of naltrexone on alcohol’s stimulant properties and self-administration behavior in social drinkers: influence of gender and genotype. Alcohol Clin Exp Res (2011) 35:1134– 1141.

175. Ralevski E, Balachandra K, Gueorguieva R, Limoncelli D, Petrakis I. Effects of Naltrexone on Cognition in a Treatment Study of Patients with Schizophrenia and Comorbid Alcohol Dependence. J Dual Diagn (2006) 2:53–69.

176. Jaffe A, O’Malley SS, Hickcox M, Chang G, Schottenfeld RS, Rounsaville BJ. Effects of Naltrexone on the Cognitive Functioning of Recently Abstinent Alcohol Dependent Patients. Novel Pharmacological Interventions for Alcoholism. Springer New York (1992). p. 329–334

177. Fletcher PJ, Tampakeras M, Sinyard J, Higgins GA. Opposing effects of 5-HT(2A) and 5-HT(2C) receptor antagonists in the rat and mouse on premature responding in the five-choice serial reaction time test. Psychopharmacology (2007) 195:223–234.

178. Lalonde R. The neurobiological basis of spontaneous alternation. Neurosci Biobehav Rev (2002) 26:91–104.

179. Floresco SB, Block AE, Tse MTL. Inactivation of the medial prefrontal cortex of the rat impairs strategy set-shifting, but not reversal learning, using a novel, automated procedure. Behav Brain Res (2008) 190:85–96.

180. Irimia C, Wiskerke J, Natividad LA, Polis IY, de Vries TJ, Pattij T, Parsons LH. Increased impulsivity in rats as a result of repeated cycles of alcohol intoxication and abstinence. Addict Biol (2015) 20:263–274.

181. Prevot TD, Li G, Vidojevic A, Misquitta KA, Fee C, Santrac A, Knutson DE, Stephen MR, Kodali R, Zahn NM, et al. Novel Benzodiazepine-Like Ligands with Various Anxiolytic, Antidepressant, or Pro-Cognitive Profiles. Mol Neuropsychiatry (2019) 5:84– 97.

182. Fletcher PJ, Rizos Z, Noble K, Higgins GA. Impulsive action induced by amphetamine, cocaine and MK801 is reduced by 5-HT(2C) receptor stimulation and 5-HT(2A) receptor blockade. Neuropharmacology (2011) 61:468–477.

183. Aharonovich E, Hasin DS, Brooks AC, Liu X, Bisaga A, Nunes EV. Cognitive deficits predict low treatment retention in cocaine dependent patients. Drug Alcohol Depend (2006) 81:313–322.

184. Aharonovich E, Nunes E, Hasin D. Cognitive impairment, retention and abstinence among cocaine abusers in cognitive-behavioral treatment. Drug Alcohol Depend (2003) 71:207–211.

185. Copersino ML, Schretlen DJ, Fitzmaurice GM, Lukas SE, Faberman J, Sokoloff J, Weiss RD. Effects of cognitive impairment on substance abuse treatment attendance: predictive validation of a brief cognitive screening measure. Am J Drug Alcohol Abuse (2012) 38:246– 250.

186. Allen DN, Goldstein G, Seaton BE. Cognitive rehabilitation of chronic alcohol abusers. Neuropsychol Rev (1997) 7:21–39.

187. Katz EC, King SD, Schwartz RP, Weintraub E, Barksdale W, Robinson R, Brown BS. Cognitive ability as a factor in engagement in drug abuse treatment. Am J Drug Alcohol Abuse (2005) 31:359–369.

188. Curran GM, Sullivan G, Williams K, Han X, Collins K, Keys J, Kotrla KJ. Emergency department use of persons with comorbid psychiatric and substance abuse disorders. Ann Emerg Med (2003) 41:659–667.

189. Dawson H, Zinck G. CIHI Survey: ED spending in Canada: a focus on the cost of patients waiting for access to an in-patient bed in Ontario. Healthc Q (2009) 12:25–28.

190. Saunders CE. Time study of patient movement through the emergency department: sources of delay in relation to patient acuity. Ann Emerg Med (1987) 16:1244–1248.

191. Rehm J, Gmel GE Sr, Gmel G, Hasan OSM, Imtiaz S, Popova S, Probst C, Roerecke M, Room R, Samokhvalov AV, et al. The relationship between different dimensions of alcohol use and the burden of disease-an update. Addiction (2017) 112:968–1001.

192. Stockwell T. Canadian Substance Use Costs and Harms in the Provinces and Territories: (2007-2014). Canadian Centre on Substance Abuse (2018). 42 p.

193. Severtson SG, von Thomsen S, Hedden SL, Latimer W. The association between executive functioning and motivation to enter treatment among regular users of heroin and/or cocaine in Baltimore, MD. Addict Behav (2010) 35:717–720.

194. Yücel M, Lubman DI. Neurocognitive and neuroimaging evidence of behavioural dysregulation in human drug addiction: implications for diagnosis, treatment and prevention. Drug Alcohol Rev (2007) 26:33–39.

195. Bruijnen CJWH, Dijkstra BAG, Walvoort SJW, Markus W, VanDerNagel JEL, Kessels RPC, DE Jong CAJ. Prevalence of cognitive impairment in patients with substance use disorder. Drug Alcohol Rev (2019) 38:435–442.

196. Roth RM, Isquith PK, Gioia GA. “Assessment of Executive Functioning Using the Behavior Rating Inventory of Executive Function (BRIEF).,” In: Goldstein S, Naglieri JA, editors. Handbook of Executive Functioning. New York, NY: Springer New York (2014). p. 301–331

197. Andersen PK, Gill RD. Cox’s Regression Model for Counting Processes: A Large Sample Study. Mathematisch Centrum, Afdeling Mathematische Statistiek (1981). 38 p.

198. Prentice RL, Williams BJ, Peterson AV. On the regression analysis of multivariate failure time data. Biometrika (1981) 68:373–379.

199. Cai J, Schaubel DE. Marginal means/rates models for multiple type recurrent event data. Lifetime Data Anal (2004) 10:121–138.

200. Therneau TM, Grambsch PM. Modeling Survival Data: Extending the Cox Model. Springer Science & Business Media (2013). 350 p.

201. Epstein EE. “Classification of alcohol-related problems and dependence.,” In: Heather N, editor. International handbook of alcohol dependence and problems (pp. Hoboken, NJ, US: John Wiley & Sons Ltd, xvii (2001). p. 47–70

202. Houghton DC, Spratt HM, Keyser-Marcus L, Bjork JM, Neigh GN, Cunningham KA, Ramey T, Moeller FG. Behavioral and neurocognitive factors distinguishing post-traumatic stress comorbidity in substance use disorders. Transl Psychiatry (2023) 13:296.

203. Oliva V, De Prisco M, Pons-Cabrera MT, Guzmán P, Anmella G, Hidalgo-Mazzei D, Grande I, Fanelli G, Fabbri C, Serretti A, et al. Machine Learning Prediction of Comorbid Substance Use Disorders among People with Bipolar Disorder. J Clin Med Res (2022) 11: doi: 10.3390/jcm11143935

204. Long Y, Pan N, Ji S, Qin K, Chen Y, Zhang X, He M, Suo X, Yu Y, Wang S, et al. Distinct brain structural abnormalities in attention-deficit/hyperactivity disorder and substance use disorders: A comparative meta-analysis. Transl Psychiatry (2022) 12:368.

205. Schuckit MA. Comorbidity between substance use disorders and psychiatric conditions. Addiction (2006) 101 Suppl 1:76–88.

206. Regier DA, Farmer ME, Rae DS, Locke BZ, Keith SJ, Judd LL, Goodwin FK. Comorbidity of mental disorders with alcohol and other drug abuse. Results from the Epidemiologic Catchment Area (ECA) Study. JAMA (1990) 264:2511–2518.

207. Silveira S, Shah R, Nooner KB, Nagel BJ, Tapert SF, de Bellis MD, Mishra J. Impact of Childhood Trauma on Executive Function in Adolescence-Mediating Functional Brain Networks and Prediction of High-Risk Drinking. Biol Psychiatry Cogn Neurosci Neuroimaging (2020) 5:499–509.

208. Deak JD, Johnson EC. Genetics of substance use disorders: a review. Psychol Med (2021) 51:2189–2200.

209. Connor JP, Haber PS, Hall WD. Alcohol use disorders. Lancet (2016) 387:988–998.

210. Niklason GR, Rawls E, Ma S, Kummerfeld E, Maxwell AM, Brucar LR, Drossel G, Zilverstand A. Explainable machine learning analysis reveals sex and gender differences in the phenotypic and neurobiological markers of Cannabis Use Disorder. Sci Rep (2022) 12:15624.

211. Saberi Zafarghandi MB, Khanipour H, Ahmadi SM. Typology of Substance Use Disorder Based on Temperament Dimensions, Addiction Severity, and Negative Emotions. Iran J Psychiatry (2018) 13:184–190.

212. Charron E, Yu Z, Lundahl B, Silipigni J, Okifuji A, Gordon AJ, Baylis JD, White A, Carlston K, Abdullah W, et al. Cluster analysis to identify patient profiles and substance use patterns among pregnant persons with opioid use disorder. Addict Behav Rep (2023) 17:100484.

213. Drossel G, Brucar LR, Rawls E, Hendrickson TJ, Zilverstand A. Subtypes in addiction and their neurobehavioral profiles across three functional domains. Transl Psychiatry (2023) 13:127.

214. Gold AK, Otto MW. Impaired risk avoidance in bipolar disorder and substance use disorders. J Psychiatr Res (2022) 152:335–342.

215. Moss HB, Chen CM, Yi H-Y. Subtypes of alcohol dependence in a nationally representative sample. Drug Alcohol Depend (2007) 91:149–158.

216. Windle M, Scheidt DM. Alcoholic subtypes: are two sufficient? Addiction (2004) 99:1508–1519.

217. Tapert SF, Eberson-Shumate S. Alcohol and the adolescent brain: What we’ve learned and where the data are taking us. Alcohol Res (2022) 42:07.

218. Barr P, Neale Z, Chatzinakos C, Schulman J, Mullins N, Zhang J, Chorlian D, Kamarajan C, Kinreich S, Pandey A, et al. Clinical, genomic, and neurophysiological correlates of lifetime suicide attempts among individuals with alcohol dependence. Res Sq (2024) doi: 10.21203/rs.3.rs-3894892/v1

219. Hueniken K, Somé NH, Abdelhack M, Taylor G, Elton Marshall T, Wickens CM, Hamilton HA, Wells S, Felsky D. Machine Learning-Based Predictive Modeling of Anxiety and Depressive Symptoms During 8 Months of the COVID-19 Global Pandemic: Repeated Cross-sectional Survey Study. JMIR Ment Health (2021) 8:e32876.

220. Tio ES, Misztal MC, Felsky D. Evidence for the biopsychosocial model of suicide: a review of whole person modeling studies using machine learning. Front Psychiatry (2023) 14:1294666.

221. Felsky D, Cannitelli A, Pipitone J. Whole Person Modeling: a transdisciplinary approach to mental health research. Discov Ment Health (2023) 3:16.

222. Sampedro-Piquero P, Ladrón de Guevara-Miranda D, Pavón FJ, Serrano A, Suárez J, Rodríguez de Fonseca F, Santín LJ, Castilla-Ortega E. Neuroplastic and cognitive impairment in substance use disorders: a therapeutic potential of cognitive stimulation. Neurosci Biobehav Rev (2019) 106:23–48.

223. Moeller SJ, Paulus MP. Toward biomarkers of the addicted human brain: Using neuroimaging to predict relapse and sustained abstinence in substance use disorder. Prog Neuropsychopharmacol Biol Psychiatry (2018) 80:143–154.

224. Tanabe J, Regner M, Sakai J, Martinez D, Gowin J. Neuroimaging reward, craving, learning, and cognitive control in substance use disorders: review and implications for treatment. Br J Radiol (2019) 92:20180942.

